# The impact of sub-national heterogeneities in demography and epidemiology on the introduction of rubella vaccination programs in Nigeria

**DOI:** 10.1101/2024.02.18.24302994

**Authors:** Taishi Nakase, Tenley Brownwright, Oyeladun Okunromade, Abiodun Egwuenu, Oladipo Ogunbode, Bola Lawal, Kayode Akanbi, Gavin Grant, Orji O. Bassey, Melissa M. Coughlin, Bettina Bankamp, Ifedayo Adetifa, Jessica Metcalf, Matthew Ferrari

## Abstract

Rubella infection during pregnancy can result in miscarriage or infants with a constellation of birth defects known as congenital rubella syndrome (CRS). When coverage is inadequate, rubella vaccination can increase CRS cases by increasing the average age of infection. Thus, the World Health Organisation recommends that countries introducing rubella vaccine be able to vaccinate at least 80% of each birth cohort. Previous studies have focused on national-level analyses and have overlooked sub-national variation in introduction risk. We characterised the sub-national heterogeneity in rubella transmission within Nigeria and modelled local rubella vaccine introduction under different scenarios to refine the set of conditions and strategies required for safe rubella vaccine use. Across Nigeria, the basic reproduction number ranged from 2.6 to 6.2. Consequently, the conditions for safe vaccination varied across states with low-risk areas requiring coverage levels well below 80%. In high-risk settings, inadequate routine coverage needed to be supplemented by campaigns that allowed for gradual improvements in vaccination coverage over time. Understanding local heterogeneities in both short-term and long-term epidemic dynamics can permit earlier nationwide introduction of rubella vaccination and identify sub-national areas suitable for program monitoring, program improvement and campaign support.

## BACKGROUND

Rubella is a childhood viral infection transmitted by respiratory droplets. It is typically characterised by a mild fever and a rash with life-long immunity following infection. Rubella infection during the first trimester of pregnancy, however, can result in miscarriage, stillbirth, and the development of severe congenital malformations which include hearing impairments, cataracts, and heart defects, collectively referred to as congenital rubella syndrome (CRS) [1]. Infants with CRS often suffer multiple life-long disabilities, requiring complex and resource-intensive care [2]. Following the development of the highly efficacious combined measles-rubella (MR) and measles-mumps-rubella (MMR) vaccines and the widespread introduction of rubella-containing-vaccine (RCV) in national childhood immunisation programs over the last two decades, reported rubella cases have decreased substantially from 670,894 in 2000 to 10,194 in 2020 [3]. As of 2020, 173 (89%) of 194 World Health Organization (WHO) Member States have introduced routine rubella immunisation with an estimated 70% of all infants vaccinated against rubella globally [4]. Economic analyses of these rubella vaccination efforts have demonstrated that the introduction of rubella vaccine is consistently cost-effective across both developed and developing countries [5].

Since rubella vaccine is delivered as a MR combination vaccine, the introduction of rubella vaccination into a national immunisation program is typically structured to build upon ongoing measles immunisation efforts [6]. To help catalyse the introduction of the vaccine into the routine health system, a wide-age-range catch-up campaign targeting children up to 15 years of age can be implemented prior to routine rubella vaccination. The introduction of RCV is also often accompanied by follow-up campaigns designed to fill gaps in immunity and bolster vaccination rates in low coverage areas.

In the WHO Eastern Mediterranean Region (EMR) and African Region (AFR), a total of 19 countries have yet to introduce RCV and an additional 21 countries have introduced rubella vaccine between 2017 and 2022 [7]. The incidence of CRS remains high with 116 cases per 100,000 live births estimated in Africa, underscoring the potential rewards of rubella vaccination programs in the region [8]. Since 2012, Gavi, the Vaccine Alliance, has invited eligible countries to apply for support towards introducing a vaccine against rubella [9]. Despite this available funding, some countries in the EMR and AFR have yet to either meet the eligibility criteria to receive Gavi support or begin the process of introducing the MR vaccine into their routine immunisation programs. Understanding the risks and possible strategies for the introduction of rubella vaccination in these remaining countries, many of which are classified as low-income, is essential towards both regional and global rubella elimination goals [10].

Generally, mathematical models predict that intermediate coverage levels for routine rubella vaccination can raise the average age of infection and thus increase the risk of rubella in pregnancy [11,12]. This theoretical risk appears to have been borne out in some countries such as Greece [13] and Costa Rica [14], where low routine vaccination coverage preceded an increase in CRS incidence. If vaccine coverage is high enough, however, the incidence of rubella decreases across all age groups such that the expected shift in the age profile of infection no longer results poses a risk to women of reproductive age [15]. Citing this potential for increases in CRS cases when RCV coverage is too low, the WHO recommends that a coverage of at least 80% be achieved and sustained for each birth cohort through a combination of routine immunisation and campaigns [6]. Gavi requires that countries meet this minimum level of coverage to be eligible for financial and technical support towards the introduction of national rubella vaccination programs [16]. A country’s ability to introduce MR vaccine is thus often tied to this 80% coverage benchmark.

Theoretical results from previous modelling studies have indicated that coverage levels of at least 80% are required to prevent increases in CRS burden if routine childhood vaccination is the sole intervention, corroborating the WHO’s recommendation [11,17,18]. More recent studies, however, suggest that the minimum safe level of coverage is highly variable, depending on local birth rates, transmission rates, and the level of imported infections [19,20]. It has become evident that in low-to-moderate birth rate settings (i.e. <30 births per 1,000 population) with estimates of the basic reproduction number (R0) under 8 and a low risk of importations, routine immunisation coverage of 80% is a relatively conservative threshold at which RCV can be safely introduced [20,21]. With higher birth rates and R0 estimates as great as 11.8 (e.g. Addis Ababa, Ethiopia [17]), the risks of suboptimal rubella vaccination in the African region are often projected to be much greater with safe levels of coverage thought to be well above the 80% threshold [20].

Past analyses of the complex interaction between the age profile of infection and nonlinear epidemic dynamics have largely focused on static vaccination strategies at equilibrium conditions, measuring the effect of vaccination on CRS burden many decades in the future [18,20]. In general, increases in CRS cases under conditions of insufficient RCV coverage (i.e. insufficient to maintain safe rubella vaccine use) do not manifest for many years. Thus, a reliance on these studies to determine safe coverage levels and inform decisions on RCV introduction can result in conservative recommendations that do not consider the potential for improvements in coverage over time. This can leave countries trapped in a state of prolonged non-introduction.

Furthermore, much of the analysis on RCV introduction has been at a national level, where minimum safe levels of coverage are defined for a country at large without consideration of sub-national variation in the risks of introduction [15,22]. This focus on national-level analyses can also contribute to overly conservative decision-making, whereby risks estimated at the national level preclude the potential benefit of local RCV introduction. The countries yet to introduce routine rubella immunisation are low-middle or low-income with limited public health resources to establish and maintain the high levels of coverage required nationally under conservative recommendations. In recent years, the WHO has called for locally tailored, sub-national solutions to achieve MR goals [23]. These discussions have focused on sub-national follow-up campaigns to fill gaps in existing routine vaccination and have not fully addressed the potential for RCV introduction programs designed according to information on sub-national variation in demography and epidemiology. It is thus of considerable interest to develop strategies for introducing MR vaccination and enhancing MR monitoring (e.g. surveillance) in different sub-national areas in ways that both prevent communities with more attainable safe levels of coverage from being held back by higher-risk communities and account for the risks that are shared across communities.

We perform our analyses on Nigeria, one of the 19 countries that has not yet established routine rubella vaccination as of the end of 2022 [24]. Nigeria presents an ideal case study due to its highly heterogeneous demographic and epidemiological conditions which largely encompass those observed across much of the African region. Understanding its epidemiological features and exploring the implications of different rubella vaccination strategies can provide lessons for the introduction of RCV in other low-income countries in the region. Furthermore, a recent nationally representative serological survey conducted in Nigeria provides a unique opportunity to quantify local variation in rubella transmission rates and to use these estimates to better inform epidemic models.

To quantify how CRS burden changes following RCV introduction in each of the thirty-six states of Nigeria and the Federal Capital Territory (FCT), we estimate local age-specific seroprevalence and force of infection, and then incorporate these estimates into a dynamic transmission model. We first explore the introduction of routine vaccination alone and solve for the cumulative rate of CRS over the 30 years as a function of coverage. This allows us to quantify local variability in the risks of introduction, the interpretation of which may be otherwise obscured by the inclusion of catch-up or follow-up campaigns. We then consider how this state-level risk may be mitigated through strengthening of routine coverage and the short-term impact of supplemental immunisation activities (SIAs) at the time of RCV introduction. Given the potentially higher risks associated with routine vaccination alone and the importance of strengthening routine coverage for rubella control to be sustainable [25], understanding the boundaries of safe RCV introduction and identifying where and how resources can be most effectively allocated to short-term interventions, such as follow-up campaigns, are of considerable public health relevance.

In this analysis, we ask three questions: 1) what is the spatial heterogeneity in transmission risk and demographic conditions across Nigeria? 2) how does sub-national heterogeneity influence the relative risks of RCV rollout across different areas in Nigeria? 3) what are the spatio-temporal consequences of RCV rollout at potentially insufficient coverage levels when we allow for improvements in coverage over time and introduce supplemental immunisation campaigns? In this way, we characterise sub-national heterogeneities in transmission rates and their implications on vaccination strategies in Nigeria and, crucially, also examine some of the conceptual barriers to the safe introduction of RCV in many developing countries without existing rubella immunisation programs.

## METHODS AND MATERIALS

### Serological survey data

The 2018 Nigeria HIV/AIDS Indicator and Impact Survey (NAIIS) collected dried blood spots from 140,000 adults aged 15-64 years and 31,459 children aged 0-14 years using a cluster randomised design across 37 state-level strata. A subset of the adult (9,737) and all the children (31,459) samples were subsequently analysed by Nigeria Centers for Disease Control (NCDC) in partnership with US Centers for Disease Control (USCDC) to determine the prevalence of IgG antibodies to rubella virus using a Luminex^®^ based multiplex bead assay [26,27]. The resulting data set included 41,099 individual records with rubella IgG status (binary: positive/negative) and age in years from 3,871 cluster locations. Individual records were aggregated to the state level to derive state-level estimates of the age-specific seroprevalence for rubella in each state (**figure S1**).

### Estimation of the force of infection and the basic reproduction number

The age-specific force of infection function for each state was estimated using the state-level age-stratified serological survey data. We used a discrete age-structured model, where the force of infection was assumed to be constant within three discrete age classes: 0-3 years, 3-15 years, and 15+ years. These age classes were selected to strike a balance between important epidemiological features of rubella (e.g. higher rates of transmission among children) and identifiability of the model parameters. Under the assumptions of life-long immunity and a constant rate of loss of maternally acquired passive immunity in infants, we derived an expression of the proportion of seropositive individuals at a given age (**equation S2**) and fitted it to the age-stratified serological survey data for each state (**figure S1**). We fitted the model using Markov Chain Monte Carlo (MCMC) methods in RStan [28] and weakly informative priors on the coefficients (**table S1**). Three chains were initialised and run for 4,000 iterations with the first 2,000 treated as warmup. Convergence and sufficient chain mixing were verified based on visualisations and the R-hat convergence diagnostic [29]. We obtained estimates of the 3x3 who-acquires-infection-from-whom (WAIFW) matrix by solving a set of equations for the piecewise constant age-specific force of infection function (**equation S10**; **figure S2**). The R0 for each state was then estimated by calculating the dominant eigenvalue of the resultant next generation matrix taken at disease-free equilibrium [30,31]. Further details including equations are provided in **Supplementary Text**.

### Dynamic transmission model

We developed a deterministic age structured MSEIRV (maternally immune, susceptible, exposed, infected, recovered, and vaccinated) compartment model to investigate the relative risks of RCV rollout at the sub-national level across Nigeria (**figure S3**). Each population was stratified into 73 age groups (monthly age strata from 0 to 4 years; yearly age strata from 4 to 20 years; 5-yearly age strata from 20 to 60 years; 60+ years age stratum) with constant rates of movement between age strata. We assumed a piecewise constant mortality rate function parameterized using published survival data [32]. Birth rates and age-specific fertility rates were set according to national census data [33]. Birth rates were assumed to decline at a rate of 1% per year to reflect expected demographic changes [32]. The loss of maternally acquired passive immunity for rubella was modelled by a constant exponential decay parameter, which was estimated from the age-stratified serological survey data. The age-specific transmission rates were described with a WAIFW matrix and derived from the estimated piecewise constant force of infection functions of each state. Seasonal forcing was incorporated into the model by varying the intensity of transmission over the year according to a cosine function (**equation S23**). The average latent period and the average infectious period were both set to 10 days and were assumed to be the same across age groups [6]. To capture the effects of inter-state travel, we assumed a relatively high yearly emigration rate of 10 infectives per 100,000 individuals. Full details on the mathematical formulation and parameterization of the model can be found in **Supplementary Text**.

The population age structure of each state was initialised according to national census data [34] with individuals partitioned into epidemiological categories based on the posterior estimates of seroprevalence derived from the serological survey data. Routine rubella vaccination was assumed to occur in infants at 9 months of age with perfect efficacy. SIAs including starting (‘catch-up’) campaigns and regular (‘follow-up’) campaigns were modelled as pulsed campaigns reaching the specified proportion of the targeted age range at the start of the year, regardless of the level of coverage attained by earlier vaccination programs.

To explore the relative risks of RCV introduction across Nigeria, we considered several different immunisation strategies. We first considered the introduction of routine vaccination of infants only at coverage levels set according to current state-level measles-containing-vaccine first-dose (MCV1) coverage [33]. Routine vaccination is seldom used alone, but this analysis served to illustrate the effect of local differences in birth rate, transmission rate and vaccination coverage on CRS incidence and the potential risks of rubella vaccination. To evaluate the appropriateness of the widely cited 80% coverage guideline at the sub-national level in Nigeria, we also identified the minimum routine vaccination coverage necessary to prevent increases in 30-year CRS burden following RCV rollout using nonlinear constrained optimization methods. We then allowed for modest improvements in routine vaccination coverage over time starting at the current routine measles coverage in each state. Specifically, we considered four yearly rates of improvement - 1%, 2.5%, 5% and 10% - where, for example, a yearly rate of improvement of 5% means that the proportion of infants who are missed by routine immunisation is decreased by 5% every year. Improvements in routine coverage were modelled in this way to reflect the fact that each percentage increase in coverage becomes more difficult as coverage increases. Finally, we expanded the model to include a catch-up campaign during the first year of rubella vaccination targeting 90% of children aged 9 months and 14 years old as well as follow-up campaigns for 90% of children aged 9 months through 4 years olds at 4-year intervals. These introduction scenarios were selected to reflect the general framework for RCV rollout adopted in the past and currently recommended by the WHO [6].

### Simulations, uncertainty propagation and comparison

We performed deterministic simulations for each scenario in each state over 30 years. The uncertainty in our estimates of the force of infection and basic reproduction number were propagated in our dynamic transmission model by repeatedly sampling from the posterior distribution of the force of infection function and performing numerous simulations for each scenario (200 for non-improvement scenarios, 100 for improvement scenarios with and without SIAs).

To track the CRS dynamics, we introduced additional pregnancy compartments in the model (**figure S3**). For each age group, we added three susceptible-and-pregnant compartments to reflect the trimesters of pregnancy as well as an exposed-and-pregnant compartment for women infected with rubella during the first trimester. Individuals were modelled to become pregnant according to state-level age-specific fertility rates with the probability of CRS following infection during the first trimester taken to be 0.65 [21]. We assessed the transient effects of the introduction of rubella vaccination by calculating the yearly CRS incidence and measured 30-year CRS burden as the ratio of total number of CRS cases and the total number of live births over 30 years.

## RESULTS

### Rubella epidemiology

Using age-stratified serological survey data collected for each state in Nigeria, we derived state-level estimates for the force of infection and basic reproduction number (figure 1). Mean estimates for R0 ranged from as low as 2.6 (90% credible interval (CI): 2.4 to 3.0) in Cross River to as high as 6.2 (90% CI: 5.3, 7.9) in Jigawa, with most states between 3 and 5 (figure 1a). The 90% credible intervals were wide for several states, with the upper limit reaching greater than 8 in Jigawa and Kano. The spatial heterogeneity in R0 followed a North-South gradient with higher estimates for states in the North than in the South (figure 1b). When the same analysis was performed at the national level on the aggregated serological survey data, the mean R0 was estimated to be 3.3 (90% CI: 3.2, 3.4). The 90% credible intervals for 24% of the states (9/37) had no overlap with that estimated at the national level.

**Figure 1.**
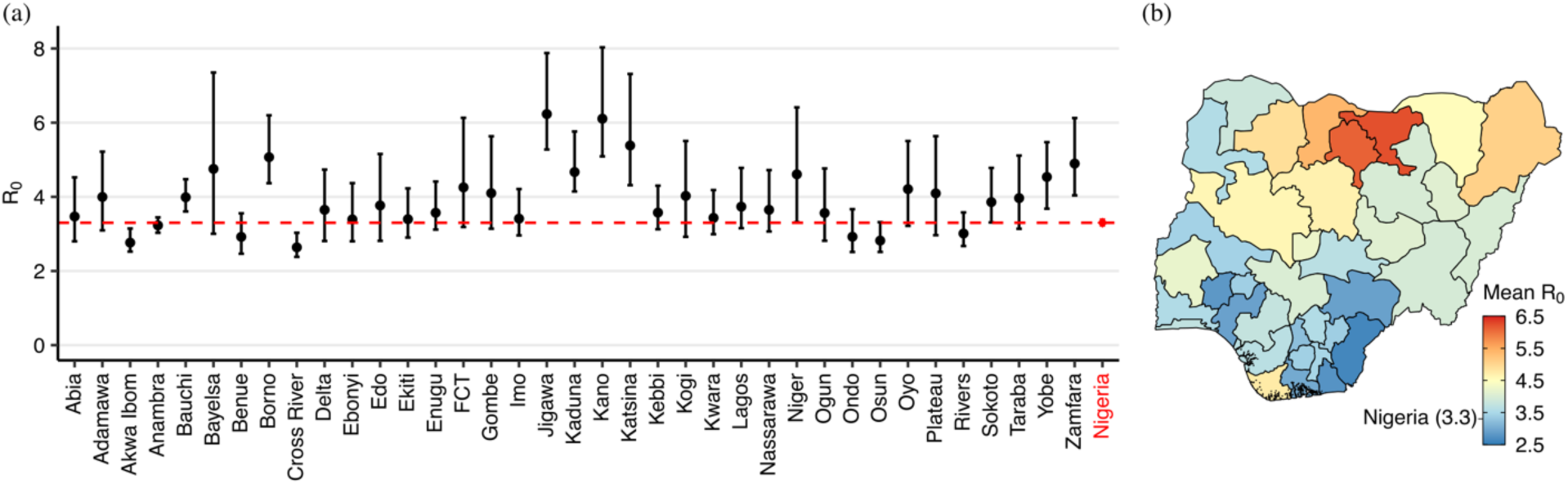
Estimates of basic reproduction number across Nigeria. **(a)** The mean estimate and the 90% credible interval of the basic reproductive number (R0) in each state (black) and at the national level (red). **(b)** Map of the mean estimate of R0 in each state and nationally.

The age-dependent risk of infection varied across states such that the mean age of infection ranged from 3.6 years (90% CI: 3.3, 3.9 years) in Jigawa to 11.9 years (90% CI: 9.8, 15 years) in Cross River (**figure S4a**). The force of infection was estimated to be highest among individuals in the 3-15 years age class, followed by individuals in the 0-3 years and 15+ years age classes (**figure S4b**). These sub-national differences in age-dependent transmission rates resulted in differences in the risk of infection among women of reproductive age. The percentage of women of childbearing age (15-44 years) estimated to have no past rubella infection and thus at risk was highly variable ranging from 0.34% (90% CI: 0.14%, 0.64%) in Jigawa to 9.9% (90% CI: 7.8%, 12%) in Cross River (**figure S5a**). Using state-level demographic information, we also estimated that the mean number of pregnant women at risk of rubella infection (i.e. no past infections) per year across Nigeria was 272,991 (90% CI: 256,429, 289,823) with the largest at-risk populations in states within the South South and South West administrative zones (**figure S5b**).

### Static analysis: routine infant vaccination only

To examine the effect of spatial heterogeneities in demographic conditions and transmission rates on the risk of RCV rollout, we projected the population in each state forwards through time and calculated the expected 30-year CRS burden with and without routine infant vaccination at current routine measles coverage (figure 2). In the absence of rubella vaccination, the spatial distribution of CRS incidence was highly heterogeneous and largely followed a North-South gradient with estimates reaching greater than 80 cases per 100,000 live births in the south and falling below 10 cases per 100,000 live births in the north (figure 2b; **figure S6**). States with higher rubella transmission rates generally had smaller current CRS burden but were expected to experience greater relative increases in infections among women of childbearing age and thus a higher relative CRS burden following the introduction of routine MR vaccination (figure 1b; figure 2c-d). For many states in the North Central, South East, South South and South West zones (e.g. Benue and Ogun) where estimated transmission is relatively low and current CRS burden is predicted to be high, coverage levels of less than 80% were sufficient to reduce 30-year CRS burden. Due to a combination of low routine coverage levels and high transmission, all states in the North East and North West zones except Adamawa were expected to have increases in CRS burden over 30 years following the introduction of RCV at current coverage levels without follow-up campaigns. CRS burden was estimated to increase by as much as 82% in Jigawa (figure 2c). When the analyses were performed at the national level with the current routine measles coverage of 54%, we projected that CRS burden would decrease by an average of 34% over thirty years (figure 2c).

**Figure 2.**
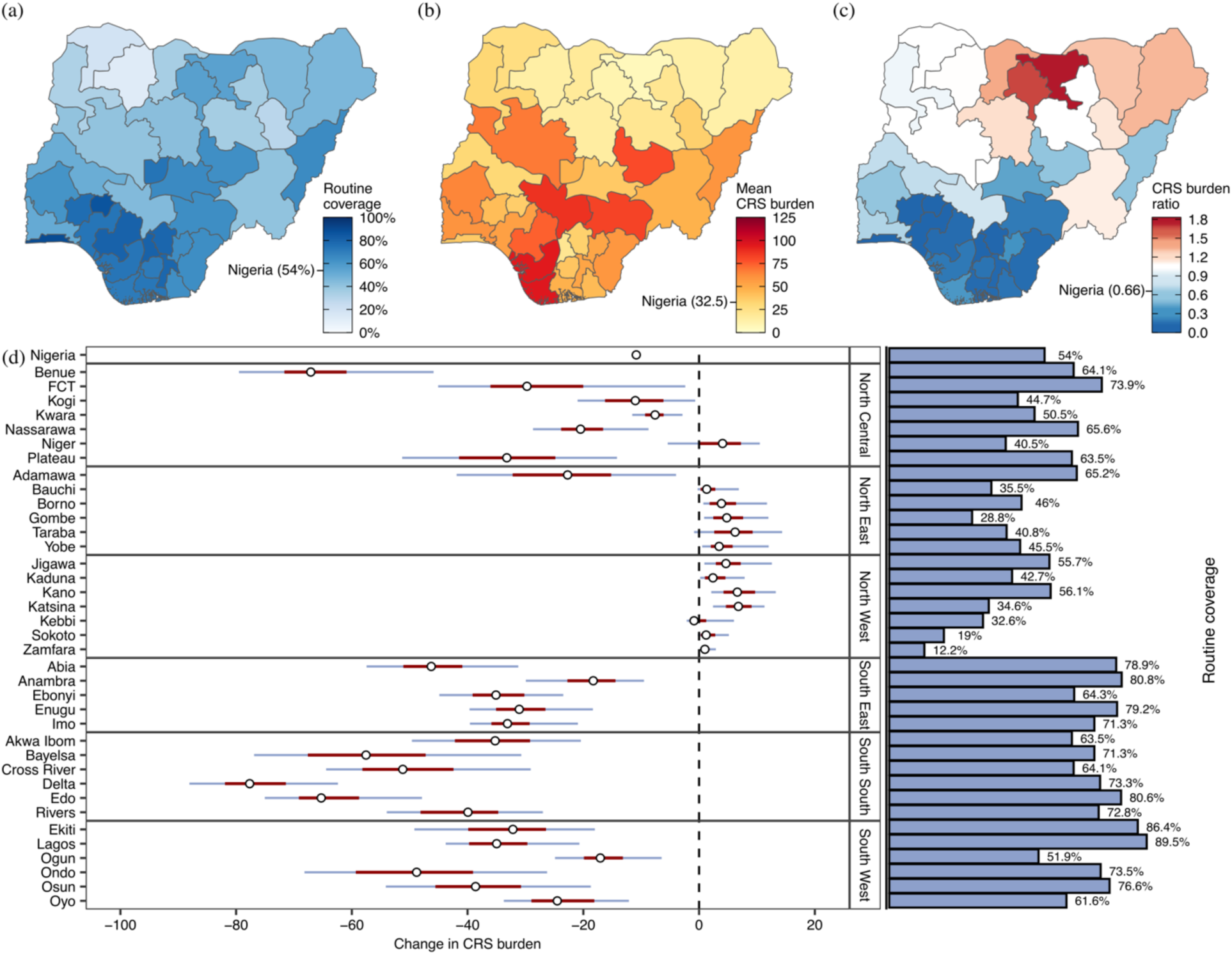
30-year CRS burden before and after introduction of routine rubella vaccination at current routine measles coverage. **(a)** Current routine measles coverage (%) in each state and at the national level. **(b)** Mean estimate of 30-year CRS burden (CRS cases per 100,000 live births) in each state and at the national level in the absence of any rubella vaccination. **(c)** Mean ratio of the 30-year CRS burden with and without routine vaccination where a ratio exceeding 1 indicates a predicted increase in CRS burden following the introduction of rubella vaccination relative to pre-vaccination burden. **(d)** Distribution of the change in 30-year CRS burden following introduction of routine rubella vaccination at current routine measles coverage in each state and at the national level (point=median; red bar=interquartile range; blue bar=90% credible interval).

We also calculated the minimum level of routine coverage required to prevent an increase in 30-year CRS burden following routine RCV introduction (without campaigns) to illustrate the diverse landscape for safe rubella vaccination (figure 3; **table S2**). At the national level, Nigeria was estimated to need a coverage level of 9% (90% CI: 0%, 14%) which is well below the current MCV1 coverage at 54%. In 68% of states (25/37), the minimum necessary coverage was also below the current measles coverage (figure 3b). In the southern zones, both birth rates and transmission rates are sufficiently low in many states (e.g. Abia and Cross River) such that the introduction of rubella vaccination was projected to not increase 30-year CRS burden even at very low coverage (figure 3a). In the North West and the North East, where transmission rates and birth rates are generally higher, minimum levels of routine coverage were often greater than current measles coverage (i.e. existing coverage was “insufficient”), reaching as high as 76% (90% CI: 69%, 84%) in Kano (figure 3a). These differences in current measles coverage and the minimum necessary coverage could be substantial; for example, it was estimated that Zamfara would need to more than quadruple its measles coverage from 12% to 68% (90% CI: 59%, 78%) in order to safely incorporate RCV into its current measles vaccination schedule in the absence of any campaigns. In several high-transmission states such as Kano and Zamfara, insufficient vaccination coverage could result in a doubling of 30-year CRS burden relative to pre-vaccination conditions (figure 3c). While the minimum necessary coverage ranged from 0% to 76% (figure 3a), the coverage required to reduce CRS incidence to below 1 case per 100,000 live births (i.e. elimination) after 30 years of vaccination was far less heterogeneous in the range of 68%-100% (**figure S7**).

**Figure 3.**
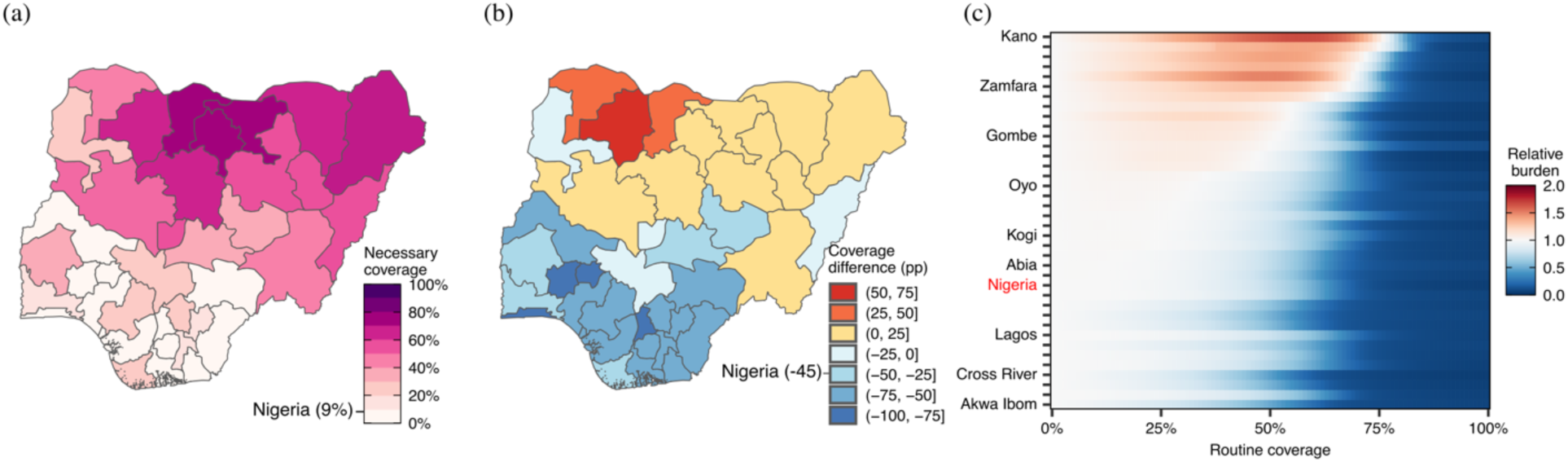
Minimum level of routine RCV coverage (without campaigns) to prevent an increase in 30-year CRS burden. **(a)** Estimated minimum necessary routine vaccination coverage in each state without campaigns. **(b)** Difference (percentage points [pp]) in the estimated minimum necessary routine coverage and the current first-dose measles-containing vaccine (MCV1) coverage where a coverage difference of 25pp means that the minimum necessary routine coverage is 25 percentage points greater than the current MCV1 coverage. **(c)** The relative 30-year CRS burden to non-vaccination projections for increasing levels of routine coverage where a value of 1.5 indicates that there is a 50% expected increase in CRS burden over the thirty years.

### Dynamic analysis: improvements in routine coverage only

Next, we simulated the introduction of routine rubella vaccination at current measles coverage and explored the effect of different rates of improvement in routine coverage over time in the absence of campaigns (figure 4; **figure S8**). In several states where current routine measles coverage was predicted to be below the minimum necessary coverage for safe rubella vaccine use (e.g. Bauchi, Borno, Kebbi, Niger), relatively modest yearly increases in routine coverage (i.e. between 1-2.5% per year) resulted in reductions in the 30-year CRS burden relative to pre-vaccination burden (figure 4a-b). For Jigawa, Kano and Katsina, where the minimum necessary RCV coverage was estimated to be upwards of 70% and the required increase in current routine coverage was greater than 25 percentage points, the yearly rate of improvement in routine coverage needed to be at least 5% for there to be a relative reduction in 30-year CRS burden (figure 4a-b).

**Figure 4.**
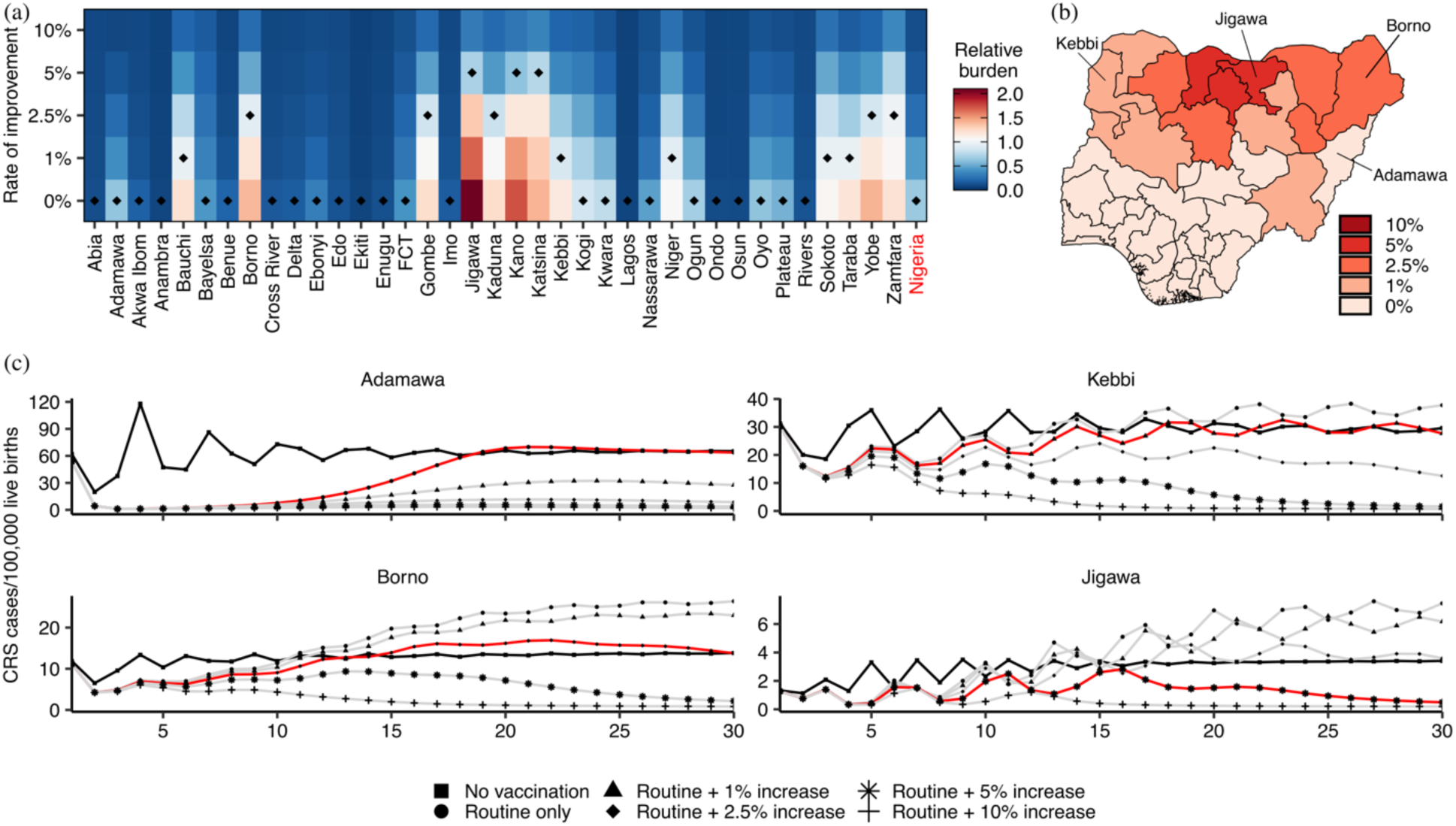
Change in CRS burden relative to pre-vaccination burden following introduction of routine RCV vaccination with different rates of improvement in coverage. **(a)** The relative 30-year CRS burden to non-vaccination projections for different yearly rates of improvement in routine coverage without follow-up campaigns. The minimum necessary rate of improvement to have a relative reduction in 30-year CRS burden for each state is indicated by a black triangle. **(b)** Minimum necessary rate of improvement in routine coverage in each state. **(c)** The yearly CRS incidence estimated for the median R0 in selected states (black=no vaccination simulation; red=simulation for the minimum necessary rate of improvement in each state; grey=other simulations). These states were selected because they are representative of the variation in rubella vaccine introduction risk across states in Nigeria (i.e. the yearly rate of improvement in routine coverage required to reduce 30-year CRS burden for Adamawa, Kebbi, Borno and Jigawa were 0%, 1%, 2.5% and 5%, respectively).

To investigate the transient effects of rubella vaccination on yearly CRS incidence across the different improvement scenarios, we calculated the number of years CRS incidence was greater in the introduction scenarios than in the non-introduction scenario and the relative difference in the CRS incidence for those years (**figure S9**). In some states decreases in 30-year CRS burden were accompanied by several years in which CRS incidence was greater than it would have been had RCV not been introduced. For example, if routine RCV vaccination were introduced at current routine measles coverage with a 2.5% yearly rate of improvement, Borno, whilst projected to have a relative reduction in 30-year CRS burden, was expected to have a median of 10 (IQR: 6, 18) years where there are a median of 2.3 (IQR: 1.3, 4.7) more CRS cases per 100,000 live births than the non-introduction scenario (figure 4c**; figure S9**). A number of other states in the North East and North West zones such as Bauchi, Gombe and Zamfara also showed long periods of higher relative CRS burden despite an expected decrease in 30-year CRS burden (**figure S8-S9**). In most states, however, relatively modest rates of improvement in routine coverage were sufficient to ensure that CRS burden decreased over 30 years without transient periods of elevated CRS incidence even when RCV was introduced at initially insufficient coverage.

### Dynamic analysis: improvements in routine coverage with campaigns

To better capture how rubella vaccination might be introduced in practice, we combined routine infant vaccination with two generalised supplemental campaign scenarios: (1) a wide-age-range catch-up campaign in the first year of vaccination targeting children aged 9 months through 14 years old only and (2) the same catch-up campaign along with follow-up campaigns in children aged 9 months through 4 years old (figure 5**; figure S10**). In all states except Jigawa and Kano, the inclusion of a catch-up campaign sufficiently reduced the pool of susceptible individuals such that routine rubella vaccination could be safely introduced without any improvements in coverage over time (**figure S11**).

**Figure 5.**
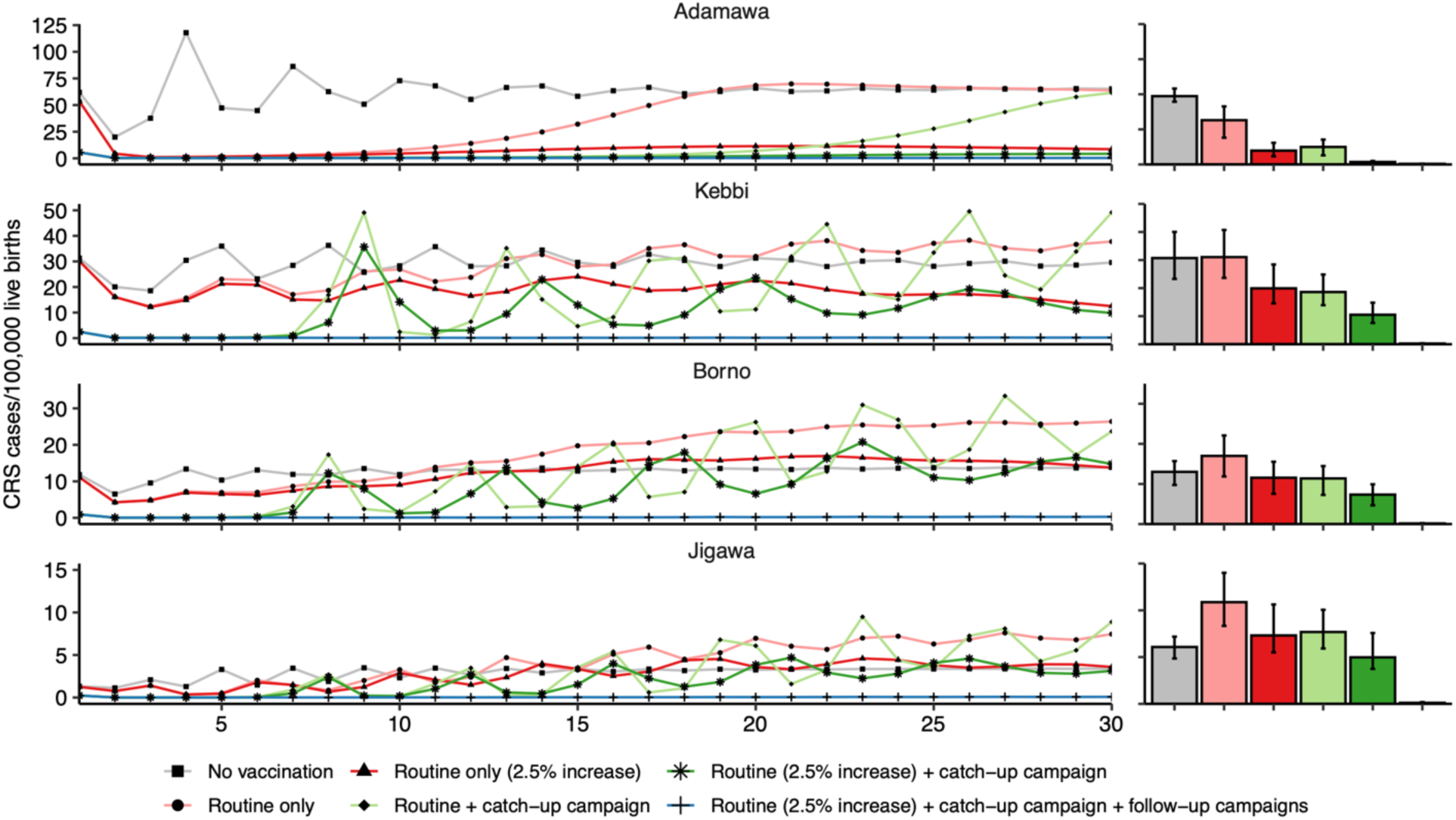
Yearly CRS incidence time series and 30-year CRS burden under different vaccination scenarios. The bar chart shows the distribution (median and interquartile range) of 30-year CRS burden under each scenario in selected states with different risks in rubella vaccine introduction. Note that the Y-axis scale is different for the panel of each state.

In 14 of these 35 states, however, the initial “honeymoon” period (i.e. the transient period of low CRS incidence after the start of rubella vaccination [35]) afforded by the catch-up campaign was short-lived with CRS incidence reaching or exceeding that expected in the absence of any vaccination towards the end of the 30 years (see light green curve for Kebbi and Borno, figure 5**; figure S10**). Critically, this rebound in CRS incidence could be prevented or greatly reduced when the routine vaccination coverage improved at a modest rate of 2.5% per year in most of these states. This combination of a wide-age-range catch-up campaign and modest yearly improvements in routine coverage was especially effective at minimising a resurgence in CRS cases in states such as Adamawa and Kebbi where the current measles coverage is close to the estimated minimum necessary coverage for safe RCV introduction (figure 5). In higher transmission states such as Katsina and Jigawa, modest improvements in routine coverage had only a marginal effect on the resurgence of CRS incidence (**figure S10**). Follow-up campaigns every 4 years virtually eliminated CRS across all states when included alongside efforts to improve routine RCV coverage over time (**figure S10**).

## DISCUSSION

Given the low cost and high efficacy of a single dose of RCV and the ease with which RCV can be delivered as part of existing measles vaccination programs, rubella vaccination is a highly cost-effective public health intervention that has the potential to greatly reduce both the incidence and the associated social and financial costs of severe birth defects in infants [5]. As of 2022, there remain 19 countries that are yet to introduce rubella vaccination. With low existing measles coverage and limited resources, some of these countries have yet to meet the 80% measles vaccine coverage guideline at the national level and receive Gavi funding [16]. Additionally, a central programmatic concern for rubella vaccine introduction is that insufficient vaccination of children may lead to increases in the average age of infection and risk of infection among susceptible women of reproductive age. There is thus a need to evaluate introduction of RCV in these resource-limited and complex settings.

Past studies have shown that the minimum safe level of vaccination coverage is not fixed and instead is highly dependent on local conditions such as transmission rates, birth rates and population age structure [20]. Yet, analyses on the safety of rubella vaccine use are often carried out at a national level, conflating sub-national differences in the relative risks of rubella vaccination. In countries where there are significant sub-national differences in epidemiological and demographic conditions, the evaluation of rubella vaccine introduction solely at the national level can result in overly conservative decision-making that prevents vaccination in communities which are at low risk of increases in CRS cases. Given that the critical vaccination coverage required to reduce CRS incidence is lowest in low birth rate, low transmission settings where CRS burden is also greatest, it is often the case that areas with the most to gain from rubella vaccination are held back by these national targets. Approaches informed by sub-national heterogeneity offer opportunities to safely and incrementally introduce rubella vaccination in countries where national coverage targets cannot yet be met but there exist local populations that could immediately benefit from rubella vaccination. These approaches can also help identify sub-national areas that need special considerations for long-term programmatic monitoring and improvement due to their greater risk.

In our analysis, we showed that there exist meaningful differences in transmission rates and demographic conditions among states in Nigeria. We found that the basic reproduction number for rubella was highly variable with higher estimates in the North than in the South. Though the range of transmission rates is in line with estimates provided in previous work [36], this study is one of only a few studies to systematically and extensively quantify rubella transmission rates at a sub-national level across an entire country [37]. The observed spatial heterogeneity in the basic reproduction number, which would be obscured in serosurveillance activities performed at the national level, shows that the epidemiology of rubella can be highly variable within a country and reinforces the value of conducting serological surveys at the sub-national level [38].

In theoretical scenarios without any campaigns, we found substantial local differences in the relative risk of RCV introduction across Nigeria. Sub-national areas where R0 and birth rates were low could safely introduce rubella vaccination at coverage levels well below that of areas where R0 and birth rates were high, consistent with previously performed national-level comparisons [20,21]. Critically, the states with the highest estimated CRS burden were predicted to benefit most from the introduction of rubella vaccination even at coverage levels well below 80%. In states with lower estimated CRS incidence much greater coverage levels were required to introduce RCV without increasing CRS incidence. The criteria for safe RCV introduction are context-dependent and not straight-forward. Yet, careful analyses informed by detailed sub-national data can help elucidate which areas can immediately benefit from routine rubella vaccination and which areas may need more support including additional monitoring and program strengthening to prepare for safe RCV introduction. In this way, a shift in the evaluation of risk from the national level to the sub-national level could help identify opportunities for rubella vaccine use that would otherwise have been indiscernible and overlooked, as well as pinpoint areas that might benefit from additional support.

When we allowed for sufficiently large improvements in vaccination coverage over time and took advantage of the delay between increases in vaccination and the concomitant changes in the age profile of infection, we saw that it was possible to introduce rubella vaccination at initially insufficient coverage levels without a resurgence in CRS cases. The inclusion of a wide-age-range catch-up campaign further increased the length of the “honeymoon” period, allowing states to minimise and even prevent the eventual resurgence in cases with relatively modest efforts to improve routine coverage over time. In some high birth rate, high transmission areas, however, these efforts were predicted to be largely ineffective at preventing a relative increase in CRS burden. These higher-risk states required aggressive short-term interventions such as follow-up campaigns to fill immunity gaps and allow routine vaccination coverage to reach levels that prevented a rebound in CRS incidence once follow-up campaigns were phased out. Overall, a combination of locally specific yearly improvement targets and follow-up campaigns in the early years of introduction provides a framework for safe rubella vaccine use in most settings where routine measles coverage is currently deemed insufficient.

We structured our analyses to focus on routine vaccination because the benefits of low-quality campaigns are transient and long-term investment in them may be unsustainable. Though some of our results may suggest that routine rubella vaccination can be safely introduced at insufficient coverage levels if combined with campaigns, there is danger of a future resurgence in CRS cases when the SIAs are inevitably phased out if these efforts are not accompanied by sustained increases in routine coverage. This underscores the need for a careful consideration of campaigns and routine immunisation strengthening as paired tools to achieve elimination. Efforts to strengthen routine immunisation should be prioritised with the awareness that supplemental campaigns, while useful in temporarily bolstering coverage and buying time for increases in routine coverage to take effect, cannot be sustained indefinitely.

Given that rubella control programs are built upon existing measles elimination efforts [6], rubella and measles vaccination strategies are typically implemented together. Since it is more infectious and its vaccine demonstrates a slightly lower efficacy than RCV, requiring two recommended childhood doses, measles tends to be more sensitive to low coverage than rubella. The frequency and timing for MR vaccination are thus often driven by measles immunity gaps rather than by concerns about RCV coverage. Nevertheless, efforts to increase MCV coverage above 95% [39] will accelerate the increases in routine RCV coverage that are necessary to achieve long-term decreases in CRS incidence.

The generally positive situation for the introduction of RCV at initially insufficient coverage levels in these deterministic simulations should be interpreted with caution in light of the role that stochasticity is known to play in the extinction-reintroduction dynamics of rubella [19,20] and other ecologically similar pathogens [40]. Local extinction over long periods can result in the build-up of susceptible individuals in older age groups, leaving a population vulnerable to a spike in maternal infections and CRS cases following reintroduction [41]. Additionally, although our model incorporates emigration events, it considers each state in isolation such that our results may overlook the additional risk presented by heterogeneous vaccination coverage in metapopulations with cross migration [20,37]. Specifically, selective vaccination in one sub-national area could reduce the seeding of infections into a neighbouring unvaccinated population and allow susceptible build-up in older age groups. This additional risk, however, is most evident in isolated unvaccinated populations where rubella circulation is primarily maintained by imports and not in unvaccinated populations like those in Nigeria where rubella is highly endemic and largely unaffected by cross migration. Once vaccination is introduced into Nigeria and states approach local elimination, these extinction-reintroduction dynamics will likely become more important and require different strategies than those presented herein which are primarily concerned with the safe introduction of RCV rather than rubella elimination. The implications of spatially heterogeneous vaccination strategies at the sub-national level in countries approaching rubella elimination should be explored in the future in order to re-evaluate the effectiveness and safety of existing efforts. As detailed epidemiological data becomes available for more countries in the AFR, future studies can also examine the impact of these sub-national strategies on both rubella vaccine introduction and rubella elimination in epidemiologic blocks of countries with substantial cross migration (e.g. Nigeria, Mali, and Niger).

Our model makes several other assumptions. We assumed that seasonal rubella dynamics followed a simple sinusoidal pattern. More extreme seasonal forcing patterns can promote stochastic fadeout and irregular large epidemics among older age groups [42], which may increase CRS incidence. Longitudinal data on the intra-annual fluctuations in rubella transmission are needed to explore these alternative seasonal dynamics. We also assumed that the probability that an individual is vaccinated during a campaign is independent of the individual’s past vaccination history. There is evidence, however, that children who do not receive routine infant immunisation are also less likely to be vaccinated in subsequent campaigns [15,43]. By assuming that campaigns target individuals at random within the population, we may overlook the possibility of campaign inefficiencies and potentially overestimate the number of additional vaccine recipients from each subsequent campaign. Identifying the proportion of the population systematically missed by vaccination efforts and discounting coverage to reflect actual population protection is an important area of future research.

It is also worth acknowledging that the sub-national differences in epidemiological conditions and the effect of vaccination on CRS incidence does not necessitate a sub-national rollout strategy. Instead, these differences emphasise that some places deemed higher risk may need closer monitoring as part of a national rollout strategy. For example, surveillance of cases or seroprevalence could be strengthened and achievement of coverage targets could be better incentivised and monitored in areas where transmission and birth rates are high. Though our analysis was parameterized according to data collected in Nigeria, it should not be considered a script for the introduction of rubella vaccination in the country. Such a script would at the very least necessitate a local analysis of programmatic requirements and priorities for RCV introduction [5] as well as an evaluation of the funding and structure of the public health system in Nigeria [44], which is beyond the scope of this modelling exercise. Instead, this analysis offers potential strategies supported by models informed by detailed local data for safe rubella vaccine use in resource-limited and spatially heterogeneous settings.

## CONCLUSIONS

The relative risks of RCV introduction are heterogeneous across space. National-level coverage targets informed by studies on static vaccination strategies at equilibrium can be overly conservative and prolong the time until introduction. This modelling study is an effort to identify and critically evaluate conceptual barriers to the safe introduction of rubella vaccination and offer insights that can provide a more targeted approach to vaccine introduction. By carefully accounting for local differences in rubella epidemiology as well as the potential for improvements in coverage over time, it is possible to safely introduce rubella vaccination into countries like Nigeria which may require additional resources and/or time to meet the same national-level targets used to inform policies in the past and to determine funding eligibility. More broadly, we demonstrated that criteria based on long-term endemic solutions to models are often overly conservative. An evaluation of the transient dynamics prior to equilibrium can facilitate earlier introduction and help identity the investments in program monitoring, routine coverage improvement and continued supplemental campaigns that are necessary to ensure that vaccination efforts are safe and sustainable.

## Data Availability

Gridded population counts for 2020 in Nigeria at a spatial resolution of 3 arc seconds were obtained from the WorldPop database (http://dx.doi.org/10.5258/SOTON/WP00699, [34]). The number of births by age group used in the calculation of age-specific fertility rates as well as the state-level MCV1 coverage were made available by the Demographic and Health Surveys (DHS) Program in Nigeria (https://dhsprogram.com/pubs/pdf/FR359/FR359.pdf, [33]). Age-stratified mortality data for Nigeria was obtained from the United Nations, Department of Economic and Social Affairs, Population Division (https://population.un.org/wpp/, [32]). Raw seroprevalence data are unavailable due to privacy consideration as datasets include global positioning system coordinates which might enable identification of location of study subjects. Only restricted individuals had access to the datasets for analyses purposes only.

## ACKNOWLEDGEMENTS

The findings and conclusions in this paper are those of the authors and do not necessarily represent the official position of the Centers for Disease Control and Prevention.

## ETHICS STATEMENT

The study was conducted in accordance with the Declaration of Helsinki, and approved by the National Health Research Ethics Committee of Nigeria (NHREC) (protocol code NHREC/01/01/2007, date of approval: 27 August 2019) and the Human Subjects Review Board at the US Centers for Disease Control and Prevention where it was determined to be non-research as the study involved testing previously collected de-identified specimens. Written informed consent to use specimens to test for diseases other than HIV was obtained from all subjects involved in the study.

## AUTHOR CONTRIBUTIONS

TN, JM and MF conceived the project and advised on the methodology. TN performed the main analyses. All authors were involved in primary data collection and curation. TN, GG, JM and MF drafted the manuscript and all authors contributed to, reviewed and approved the final manuscript.

## DATA AND CODE AVAILABILITY

Gridded population counts for 2020 in Nigeria at a spatial resolution of 3 arc seconds were obtained from the WorldPop database [34]. The number of births by age group used in the calculation of age-specific fertility rates as well as state-level MCV1 coverage were made available by the Demographic and Health Surveys (DHS) Program in Nigeria [33]. Age-stratified mortality data for Nigeria was obtained from the United Nations, Department of Economic and Social Affairs, Population Division [32]. Raw seroprevalence data are unavailable due to privacy consideration as datasets include global positioning system coordinates which might enable identification of location of study subjects. Only restricted individuals had access to the datasets for analyses purposes only. All code used in the primary analyses are available on GitHub (https://github.com/TCNLAB24/crs_nigeria).

## SUPPLEMENTARY MATERIALS

**Supplementary Text** includes a full description of the statistical analysis of the serological survey data and the deterministic epidemic model as well as Supplementary Figures S1-S11 and Supplementary Tables S1-S2.

## REFERENCES

1. Cooper LZ, Ziring PR, Ockerse AB, Fedun BA, Kiely B, Krugman S. 1969 Rubella. Clinical manifestations and management. Am J Dis Child 118, 18–29.

2. Banatvala JE, Brown DWG. 2004 Rubella. Lancet 363, 1127–37. (doi:10.1016/S0140-6736(04)15897-2)

3. Zimmerman LA, Knapp JK, Antoni S, Grant GB, Reef SE. 2022 Progress Toward Rubella and Congenital Rubella Syndrome Control and Elimination — Worldwide, 2012–2020. MMWR Morb Mortal Wkly Rep 71, 196–201. (doi:10.15585/mmwr.mm7106a2)

4. Zimmerman LA, Knapp JK, Antoni S, Grant GB, Reef SE. 2022 Progress Toward Rubella and Congenital Rubella Syndrome Control and Elimination — Worldwide, 2012–2020. MMWR Morb Mortal Wkly Rep 71, 196–201. (doi:10.15585/mmwr.mm7106a2)

5. Hinman AR, Irons B, Lewis M, Kandola K. 2002 Economic analyses of rubella and rubella vaccines: a global review. Bull World Health Organ 80, 264–70.

6. World Health Organization. 2020 Rubella vaccines: WHO position paper – July 2020. Weekly Epidemiological Record 95, 306–324.

7. World Health Organization. In press. Rubella vaccination coverage. See https://immunizationdata.who.int/pages/coverage/RCV.html (accessed on 4 February 2024).

8. Vynnycky E et al. 2016 Using Seroprevalence and Immunisation Coverage Data to Estimate the Global Burden of Congenital Rubella Syndrome, 1996-2010: A Systematic Review. PLoS One 11, e0149160. (doi:10.1371/journal.pone.0149160)

9. Burki T. 2012 GAVI Alliance to roll out rubella vaccine. Lancet Infect Dis 12, 15–16. (doi:10.1016/S1473-3099(11)70362-0)

10. World Health Organization. 2020 Immunization Agenda 2030: A Global Strategy to Leave No One Behind.

11. Anderson RM, May RM. 1983 Vaccination against rubella and measles: quantitative investigations of different policies. J Hyg (Lond) 90, 259–325. (doi:10.1017/s002217240002893x)

12. Knox EG. 1980 Strategy for Rubella Vaccination. Int J Epidemiol 9, 13–23. (doi:10.1093/ije/9.1.13)

13. Panagiotopoulos T, Antoniadou I, Valassi-Adam E, Berger A. 1999 Increase in congenital rubella occurrence after immunisation in Greece: retrospective survey and systematic review How does herd immunity work? BMJ 319, 1462–1467. (doi:10.1136/bmj.319.7223.1462)

14. Morice A, Ulloa-Gutierrez R, Ávila-Agüero ML. 2009 Congenital rubella syndrome: progress and future challenges. Expert Rev Vaccines 8, 323–331. (doi:10.1586/14760584.8.3.323)

15. Winter AK et al. 2022 Feasibility of measles and rubella vaccination programmes for disease elimination: a modelling study. Lancet Glob Health 10, e1412–e1422. (doi:10.1016/S2214-109X(22)00335-7)

16. Gavi. 2023 Gavi Vaccine Funding Guidelines.

17. Cutts FT, Abebe A, Messele T, Dejene A, Enquselassie F, Nigatu W, Nokes DJ. 2000 Sero-epidemiology of rubella in the urban population of Addis Ababa, Ethiopia. Epidemiol Infect 124, 467–79. (doi:10.1017/s0950268899003532)

18. Gay NJ. 1998 Modeling measles, mumps, and rubella: implications for the design of vaccination programs. Infect Control Hosp Epidemiol 19, 570–3. (doi:10.1086/647875)

19. Metcalf CJE et al. 2013 Implications of spatially heterogeneous vaccination coverage for the risk of congenital rubella syndrome in South Africa. J R Soc Interface 10, 20120756. (doi:10.1098/rsif.2012.0756)

20. Metcalf CJE, Lessler J, Klepac P, Cutts F, Grenfell BT. 2012 Impact of birth rate, seasonality and transmission rate on minimum levels of coverage needed for rubella vaccination. Epidemiol Infect 140, 2290–301. (doi:10.1017/S0950268812000131)

21. Vynnycky E, Gay NJ, Cutts FT. 2003 The predicted impact of private sector MMR vaccination on the burden of Congenital Rubella Syndrome. Vaccine 21, 2708–19. (doi:10.1016/s0264-410x(03)00229-9)

22. Lessler J, Metcalf CJE. 2013 Balancing Evidence and Uncertainty when Considering Rubella Vaccine Introduction. PLoS One 8, e67639. (doi:10.1371/journal.pone.0067639)

23. World Health Organization. 2021 Meeting of the Strategic Advisory Group of Experts on Immunization, 22–24 March 2021: conclusions and recommendations/Reunion du Groupe strategique consultatif d’experts sur la vaccination, 22-24 mars 2021–conclusions et recommandations. Weekly epidemiological record 96, 197.

24. Omoleke SA, Udenenwu HC. 2016 Incidence of rubella in a state in North-western Nigeria: a call for action. Pan African Medical Journal 25. (doi:10.11604/pamj.2016.25.49.10003)

25. World Health Organization. 2018 Meeting of the Strategic Advisory Group of Experts on immunization, April 2018 - conclusions and recommendations. *Weekly Epidemiological Record 93*, 329–343.

26. Tohme RA et al. 2023 Tetanus and Diphtheria Seroprotection among Children Younger Than 15 Years in Nigeria, 2018: Who Are the Unprotected Children? Vaccines (Basel) 11, 663. (doi:10.3390/vaccines11030663)

27. Coughlin MM et al. 2021 Development of a Measles and Rubella Multiplex Bead Serological Assay for Assessing Population Immunity. J Clin Microbiol 59. (doi:10.1128/JCM.02716-20)

28. Stan Development Team. 2023 RStan: the R interface to Stan.

29. Vehtari A, Gelman A, Simpson D, Carpenter B, Bürkner P-C. 2021 Rank-Normalization, Folding, and Localization: An Improved R^ for Assessing Convergence of MCMC (with Discussion). Bayesian Anal 16. (doi:10.1214/20-BA1221)

30. Farrington CP, Kanaan MN, Gay NJ. 2001 Estimation of the Basic Reproduction Number for Infectious Diseases from Age-Stratified Serological Survey Data. J R Stat Soc Ser C Appl Stat 50, 251–292. (doi:10.1111/1467-9876.00233)

31. Diekmann O, Heesterbeek JAP, Metz JAJ. 1990 On the definition and the computation of the basic reproduction ratio R 0 in models for infectious diseases in heterogeneous populations. J Math Biol 28. (doi:10.1007/BF00178324)

32. United Nations Department of Economic and Social Affairs Population Division. 2022 World Population Prospects 2022, Online Edition.

33. National Population Commission (NPC) [Nigeria] and ICF. 2019 Nigeria Demographic and Health Survey 2018.

34. Bondarenko M, Kerr D, Sorichetta A, Tatem A, WorldPop. 2020 Estimates of total number of people per grid square, adjusted to match the corresponding UNPD 2020 estimates and broken down by gender and age groupings, produced using Ecopia.AI and Maxar Technologies building footprints. *University of Southampton*. See 10.5258/SOTON/WP00699.

35. Mclean AR, Anderson RM. 1988 Measles in developing countries. Part II. The predicted impact of mass vaccination. Epidemiol Infect 100, 419–442. (doi:10.1017/S0950268800067170)

36. Papadopoulos T, Vynnycky E. 2022 Estimates of the basic reproduction number for rubella using seroprevalence data and indicator-based approaches. PLoS Comput Biol 18, e1008858. (doi:10.1371/journal.pcbi.1008858)

37. Wesolowski A et al. 2016 Introduction of rubella-containing-vaccine to Madagascar: implications for roll-out and local elimination. J R Soc Interface 13, 20151101. (doi:10.1098/rsif.2015.1101)

38. Metcalf CJE, Farrar J, Cutts FT, Basta NE, Graham AL, Lessler J, Ferguson NM, Burke DS, Grenfell BT. 2016 Use of serological surveys to generate key insights into the changing global landscape of infectious disease. The Lancet 388, 728–730. (doi:10.1016/S0140-6736(16)30164-7)

39. World Health Organization. 2017 Measles vaccines: WHO position paper–April 2017/ Note de synthese de l’OMS sur les vaccins contre la rougeole–Avril 2017. Weekly Epidemiological Record 92, 205.

40. Grenfell BT, Bjørnstad ON, Kappey J. 2001 Travelling waves and spatial hierarchies in measles epidemics. Nature 414, 716–723. (doi:10.1038/414716a)

41. Metcalf CJE, Munayco C V, Chowell G, Grenfell BT, Bjørnstad ON. 2011 Rubella metapopulation dynamics and importance of spatial coupling to the risk of congenital rubella syndrome in Peru. J R Soc Interface 8, 369–76. (doi:10.1098/rsif.2010.0320)

42. Ferrari MJ, Grais RF, Bharti N, Conlan AJK, Bjørnstad ON, Wolfson LJ, Guerin PJ, Djibo A, Grenfell BT. 2008 The dynamics of measles in sub-Saharan Africa. Nature 451, 679–684. (doi:10.1038/nature06509)

43. Lessler J, Metcalf CJE, Grais RF, Luquero FJ, Cummings DAT, Grenfell BT. 2011 Measuring the Performance of Vaccination Programs Using Cross-Sectional Surveys: A Likelihood Framework and Retrospective Analysis. PLoS Med 8, e1001110. (doi:10.1371/journal.pmed.1001110)

44. Ophori EA, Tula MY, Azih A V., Okojie R, Ikpo PE. 2014 Current Trends of Immunization in Nigeria: Prospect and Challenges. Trop Med Health 42, 67–75. (doi:10.2149/tmh.2013-13)

