## Supplement Text for "The impact of sub-national heterogeneities in demography and epidemiology on the introduction of rubella vaccination programs in Nigeria"

Nakase *et al.*

**2 Supplementary figures 9**

**3 Supplementary tables 28**

### 1 Supplementary methods

#### 1.1 Analysis of age-stratified serological data

##### 1.1.1 Equations for the proportion susceptible

The population of each state $s \in S$ was structured into three discrete age classes $[a_{i-1}, a_i)$ for $i \in \{1, 2, 3\}$ (0-3 years, 3-15 years and 15+ years) within which the average force of infection $\lambda_{(s,i)}$ was assumed to be constant. In other words, we assumed a piecewise constant force of infection function of the following form for each state:

$$\lambda_s(a) = \begin{cases} \lambda_{(s,1)}, a \in [0, 3) \\ \lambda_{(s,2)}, a \in [3, 15) \\ \lambda_{(s,3)}, a \in [15, \infty) \end{cases} \tag{S1}$$

This age stratification was selected to reflect important epidemiological features of rubella (i.e. higher transmission rates among school-aged children) and ensure that the model parameters were identifiable. Additionally, we assumed that maternally-acquired passive immunity wanes at a constant exponential rate $\omega_s$ and that infection confers life-long permanent immunity. In a large, unvaccinated population at endemic equilibrium (ignoring seasonality), the proportion susceptible among individuals of age $a \in [a_{i-1}, a_i)$ is given by the following equation:

$$S_s(a) = \frac{\omega_s}{\omega_s - \lambda_{(s,1)}} \exp\left(-\int_0^a \lambda_s(t)dt\right) \times \left(1 + (\omega_s - \lambda_{(s,1)}) \times (f_1(s,i) - f_2(s,i))\right) \tag{S2}$$

where

$$f_1(s,i) = \left(\sum_{j=1}^{i-1} \frac{1}{\lambda_{(s,j)} - \omega_s} \exp\left(\int_0^{a_j} \lambda_s(t)dt - \omega_s a_j\right)\right) + \frac{1}{\lambda_{(s,i)} - \omega_s} \exp\left(\int_0^a \lambda_s(t)dt - \omega_s a\right) \tag{S3}$$

$$f_2(s,i) = \sum_{j=2}^{i} \frac{1}{\lambda_{(s,j)} - \omega_s} \exp\left(\int_0^{a_{j-1}} \lambda_s(t)dt - \omega_s a_{j-1}\right) \tag{S4}$$

Note that $f_1(s,1) = \frac{1}{\lambda_{(s,1)} - \omega_s} \exp\left(\lambda_{(s,1)}a - \omega_s a\right)$ and $f_2(s,1) = 0$.

##### 1.1.2 Estimation of force of infection from seroprevalence data

The equation for the proportion of susceptible individuals at age $a$, $S_s(a)$, was fitted to the seroprevalence data using Markov Chain Monte Carlo methods implemented in RStan

[1]. The observed number of seropositive individuals at age $a \in \mathbb{Z}^+$, $K_{s,a}$, was assumed to follow a binomial distribution:

$$K_{s,a} \sim \text{Bin}(N_{s,a}, p_s(a; \lambda_s, \omega_s))) \tag{S5}$$

where $N_{s,a}$ is the number of tested individuals at age $a$ and $p_s(a; \lambda_s, \omega_s) = 1 - S_s(a; \lambda_s, \omega_s)$ is the seroprevalence among $a$-year-old individuals in the population.

It has been shown for several childhood infections including rubella that the duration of protection by maternal antibodies can be longer in infants whose mothers have naturally-acquired immunity as opposed to vaccine-induced immunity [2]. Given that widespread vaccination is yet to be introduced in Nigeria, it was assumed that seropositive individuals have naturally-acquired immunity such that the rate of loss of maternally-acquired passive immunity is constant across states in Nigeria where $\omega = \omega_s$ for all $s \in S$.

In Taraba and Katsina, the percentage seropositivity reaches close to 100% by age 15, resulting in divergent transitions in the Hamiltonian Monte Carlo procedure (see [3]) and identifiability issues in the estimation of $\lambda_{(s,3)}$ for these states. To overcome these identifiability issues while still maintaining the epidemiological realism of the model, we made the simplifying assumption that the force of infection for the 15+ years age group was the same between each of these states and one of their neighboring states. Taraba was paired with Adamawa and Katsina with Kano such that $\lambda_{(s,3)}$ is jointly fitted across the seroprevalence data of each pair. The pairs were selected based on geographical proximity and the similarity of their serological profiles. Since rubella is primarily a childhood infection, the averaging effect on the force of infection across these pairs of states was expected to be small and not affect the overall conclusions. All other parameters were specific to each state.

The piecewise constant force of infection function $\lambda_s$ and the rate of loss of maternally-acquired passive immunity $w$ for each state were jointly estimated by fitting all parameters to the state-level seroprevalence data. We used weakly informative priors on the force of infection parameters and the exponential decay rate of maternally-derived antibodies (table S1). Three chains were initialized and run for 4000 iterations with the first 2000 treated as warmup. Convergence and sufficient chain mixing were verified based on visualizations and the R-hat convergence diagnostic. The posterior predictive distributions for each state are provided in figure S1.

##### 1.1.3 Estimation of the who-acquires-infection-from-whom matrix

For the following, we drop the $s$ subscript since the estimation process was the same for each state. When estimating the who-acquires-infection-from-whom (WAIFW) matrix $B$, we considered the following contact structure discussed in [4]:

$$B = \begin{pmatrix} \beta_1 & \beta_1 & \beta_3 \\ \beta_1 & \beta_2 & \beta_3 \\ \beta_3 & \beta_3 & \beta_3 \end{pmatrix}$$

where, for instance, the rate at which an infected individual in age group 1 (i.e. 0-3 years) infects a susceptible individual in age group 2 (i.e. 3-15 years) is $\beta_{2,1} = \beta_1$. This rate is commonly referred to as the effective contact rate, where an effective contact is defined as one that is sufficient to result in an transmission event. This contact structure was selected to capture the higher transmission rates expected among school-aged children [5]. It was limited to three degrees of freedom so that the $B$ matrix was identifiable.

We assumed that the the force of infection at time $t$ for individuals in age group $i$ was described by the following equation:

$$\lambda_i(t) = \frac{N_i(0)}{N_i(t)} \sum_{j=1}^{3} \beta_{i,j} I_j(t) \tag{S6}$$

where $N_i(t)$ is the total number of individuals and $I_i(t)$ is the number of infected individuals in age group $i$ at time $t$, respectively. At endemic equilibrium, the equation for the force of infection simplifies to the following:

$$\lambda_i^* = \frac{N_i(0)}{N_i^*} \sum_{j=1}^{3} \beta_{i,j} I_j^* \tag{S7}$$

where $\lambda_i^*$ is the force of infection, $I_i^*$ is the number of infected individuals and $N_i^*$ is the total number of individuals in age group $i$ at endemic equilibrium. We considered a piecewise constant mortality rate function such that $\mu_i$ is constant on $(a_{i-1}, a_i]$, where $L = \int_0^\infty \exp\left(\int_0^x \mu(t)dt\right) dx$ is defined as the average life expectancy. The mortality rates were estimated from survival data for the period 2015-2020 sourced from UN population databases [6]. Since the average infectious period $D$ and average period of maternally-acquired passive immunity $1/\omega$ are short on the timescale over which $\mu(a)$ varies (i.e. days vs. years), the number of infected individuals at endemic equilibrium can be approximated as follows [7]:

$$I_j^* \approx \frac{ND}{L} \lambda_j^* \int_{a_{j-1}}^{a_j} S(y) \exp(-\int_0^y \mu(t)dt) dy \tag{S8}$$

where $S(y)$ is given by equation (S2).

We can write equation (S7) in the following matrix notion:

$$\lambda^* = P(\lambda^*)\beta \tag{S9}$$

where $\beta = (\beta_1, \beta_2, \beta_3)^T$ are the WAIFW matrix parameters and $P(\lambda^*)$ is a unique matrix. The structure of the matrix $B$ was selected such that $P(\lambda^*)$ was invertible. The parameters $\beta$ were thus estimated by solving the following equation:

$$\beta = P(\lambda^*)^{-1}\lambda^* \tag{S10}$$

##### 1.1.4 Estimation of derived quantities

**Basic reproductive number**
The $R_0$ for each state was obtained by calculating the dominant eigenvalue of the next generation matrix defined by $XY$ where $X$ is a matrix with diagonal entries

$$\int_{a_{i-1}}^{a_i} \exp\left(\int_0^x \mu(t)dt\right) dx \tag{S11}$$

and $Y$ denotes the matrix with elements $Y_{i,j} = (ND/L)B_{i,j}$ [7, 8].

**Mean age of infection**
The mean age of infection at endemic equilibrium in each state was estimated according to the following expression:

$$\int_0^{\infty} S_s(t; \hat{\lambda}_s^*, \hat{\omega})dt \tag{S12}$$

where $S_s(t; \lambda_s^*, \omega)$ is the probability that an individual is susceptible at time $t$ parameterized according to the derived estimates for the force of infection function $\hat{\lambda}_s^*$ and the rate of loss of maternally-acquired passive immunity $\hat{\omega}$.

**Number of pregnant women at risk**
The number of pregnant women at risk of infection per year was estimated by summing the product of the age-specific fertility rate $f_{s,a}$ by the number of individuals with no history of rubella infection across all age groups:

$$\sum_a f_{s,a}(1 - p_s(a; \hat{\lambda}_s^*, \hat{\omega}))N_{s,a} \tag{S13}$$

where $p_s(a; \hat{\lambda}_s^*, \hat{\omega})$ is the probability that an $a$-year-old individual has been infected in the past according to the derived estimates for the force of infection function $\hat{\lambda}_s^*$ and the rate of loss of maternally-acquired passive immunity $\hat{\omega}$.

#### 1.2 Deterministic MSEIRV compartment model

We used an age-structured MSEIRV compartment model depicted in figure S5. There were three immunity profiles (transiently fully immune from maternal antibodies - $M$;

fully susceptible - $S$, $SP^{(1)}$, $SP^{(2)}$, $SP^{(3)}$; and fully immune - $R$, $V$) and two infected classes (infected but not yet infectious - $E$, $EP$; and infected and infectious - $I$). The population of size $N$ was stratified into 73 age groups (monthly age strata from 0 to 4 years; yearly age strata from 4 to 20 years; 5-yearly age strata from 20 to 60 years; 60+ years age stratum) with epidemiological variables for each age group denoted with a subscript $a$.

Individuals enter the maternally-derived passive immunity class $M_1$ at a per-capita birth rate $b(t)$. Birth rates were assumed to decline at a rate of 1% per year such that $b(t) = 0.99^t b(0)$. Maternally-derived passive immunity wanes at an exponential rate $\omega$ after which infants enter the fully susceptible class $S$. Infections of fully susceptible individuals in age class $a$ occur at a per-capita rate of $\lambda_a = \frac{N_a(0)}{N_a(t)} \sum_j \beta_{a,j}(t) I_j$. Newly infected individuals $E$ then experience an exponentially distributed latent period with a rate parameter $\sigma$ after which they become infectious. Infectiousness individuals $I$ recover at a rate of $\gamma$ and enter a fully immune recovered class $R$. Individuals of age $a$ move into the next age group at a constant rate $\theta_a = 1/(\text{length of age group})$ and are removed from the population at an age-specific natural-mortality rate $\mu_a$.

To quantify the CRS dynamics in the population, additional pregnancy compartments - $SP^{(1)}$, $SP^{(2)}$, $SP^{(3)}$, $EP$ - were introduced into the model. Fully susceptible individuals of age $a$ become pregnant $S_a$ and enter the first-trimester susceptible compartment $SP_a^{(1)}$ according to their corresponding age-specific fertility rate $f_a(t) = 0.99^t \frac{N_a(0)}{N_a(t)} f_a(0)$. Individuals move through the three susceptible trimester compartments and return to the fully susceptible at a rate $\delta$. To track infections among women during the first trimester of their pregnancies, we used an additional infected-not-yet-infectious compartment $EP$ to which individuals in $SP_a^{(1)}$ enter at a per-capita rate $\lambda_a = \sum_j \beta_{a,j}(t) I_j$. These transitions among epidemiological classes are described by the following set of differential equations:

$$\frac{dM_a}{dt} = b_a N - (\omega + \mu_a + \omega_a)M_a + \theta_{a-1}M_{i-1} \tag{S14}$$

$$\frac{dS_a}{dt} = \omega M_a + \delta SP_a^{(3)} - (\lambda_a + \mu_a + \theta_a + f_a)S_a + \theta_{a-1}S_{a-1} \tag{S15}$$

$$\frac{dSP_a^{(1)}}{dt} = f_a S_a - (\mu_a + \theta_a + \delta + \lambda_a)SP_a^{(1)} + \theta_{i-1}SP_{a-1}^{(1)} \tag{S16}$$

$$\frac{dSP_a^{(2)}}{dt} = \delta SP_a^{(1)} - (\mu_a + \theta_a + \delta + \lambda_a)SP_a^{(2)} + \theta_{i-1}SP_{a-1}^{(2)} \tag{S17}$$

$$\frac{dSP_a^{(3)}}{dt} = \delta SP_a^{(2)} - (\mu_a + \theta_a + \delta + \lambda_a)SP_a^{(3)} + \theta_{i-1}SP_{a-1}^{(3)} \tag{S18}$$

$$\frac{dE_a}{dt} = \lambda_a(S_a + SP_a^{(2)} + SP_a^{(3)}) - (\sigma + \mu_a + \theta_a)E_a + \theta_{a-1}E_{a-1} \tag{S19}$$

$$\frac{dEP_a}{dt} = \lambda_a SP_a^{(1)} - (\sigma + \mu_a + \theta_a)EP_a + \theta_{a-1}EP_{a-1} \tag{S20}$$

$$\frac{dI_a}{dt} = \sigma(E_a + EP_a) + \epsilon N - (\gamma + \mu_a + \theta_a)I_a + \theta_{a-1}I_{a-1} \tag{S21}$$

$$\frac{dR_a}{dt} = \gamma I_a - (\mu_a + \theta_a)R_a + \theta_{a-1}R_{a-1} \tag{S22}$$

##### 1.2.1 Seasonality

Seasonality in transmission intensity was assumed to follow a sinusoidal function and was defined by the following equation:

$$\beta_{a,j}(t) = \overline{\beta_{a,j}}(1 + \alpha \cos(2\pi t/365)) \tag{S23}$$

where $\overline{\beta_{a,j}}$ is the average rate at which infected individuals in age group $j$ infect susceptible individuals in age class $j$ and $\alpha$ is the intensity of seasonality. For example, $\alpha = 0$ assumes there is no seasonality while $\alpha = 0.5$ assumes that peak and trough transmission intensity is 50% more and less than the average respectively. Seasonality was assumed to affect individuals of all age groups in the same way. For all analyses, the amplitude of the seasonal variation in transmission intensity $\alpha$ was set to 20% in line with previous analyses [9].

##### 1.2.2 External immigration

In all simulations, we allowed for the migration of infectious individuals from external populations. The immigration of infectious individuals was assumed to be permanent and defined by a per-capita rate $\epsilon$ such that the number of immigration events scales with the size of a population.

##### 1.2.3 Vaccination

Individuals could be vaccinated through either routine infant immunization programs or supplemental immunization activities (SIAs) targeted at specific age groups. It was assumed that infants receive a fully-efficacious rubella-containing vaccine (RCV) at 9 months of age. Routine vaccination was introduced into the model by modifying the aging process for individuals in the 8-9 months and 9-10 months age groups. For routine coverage of $p_v$, a proportion $(1 - p_v)$ of 9-month-olds were assumed to transition into the same epidemiological compartment of the next oldest age group (i.e. 9-10 months) while the remaining proportion $p_v$ of 9-month-olds were assumed to transition into the vaccinated compartment $V$ of the next oldest age group. In this way, we ensured that a proportion $p_v$ of 9-month-olds were vaccinated against rubella. This was introduced into the system of differential equations modifying the aging process into the 9-10 months age group (i.e. $a = 9$) in the following way:

$$\frac{dX_{10}}{dt} = \ldots + (1 - p_v)\theta_9 M_9 \tag{S24}$$

$$\frac{dV_{10}}{dt} = \ldots + p_v\theta_9(M_9 + S_9 + SP_9^{(1)} + SP_9^{(2)} + SP_9^{(3)} + EP_9 + E_9 + R_9) \tag{S25}$$

where $X$ denotes all epidemiological compartments except $I$ and $V$. SIAs were modeled as pulsed campaigns where all eligible individuals were moved from their current epidemiological compartment to the vaccinated compartment $V$ at the start of the campaign year.

#### 1.3 Nonlinear constrained optimization

The minimum routine rubella vaccination coverage necessary to prevent increases in 30-year CRS burden was estimated using nonlinear constrained optimization. Due to computational constraints, we focused on estimating the minimum necessary coverage $x$ at the median $R_0$ for each state. The objective function and constraints for each state was defined as follows:

$$\min_{x \in \mathbb{R}} \quad x \tag{S26}$$

$$\text{s.t.} \quad f(x) - f(0) \leq 0 \tag{S27}$$

$$x \leq 1 \tag{S28}$$

$$-x \leq 0 \tag{S29}$$

$f(x)$ denotes the 30-year CRS burden following introduction of routine rubella vaccination of infants at coverage $x$. The optimization problems were solved using the COBYLA algorithm [10] provided in the *nloptr* package[11] in the R programming environment.

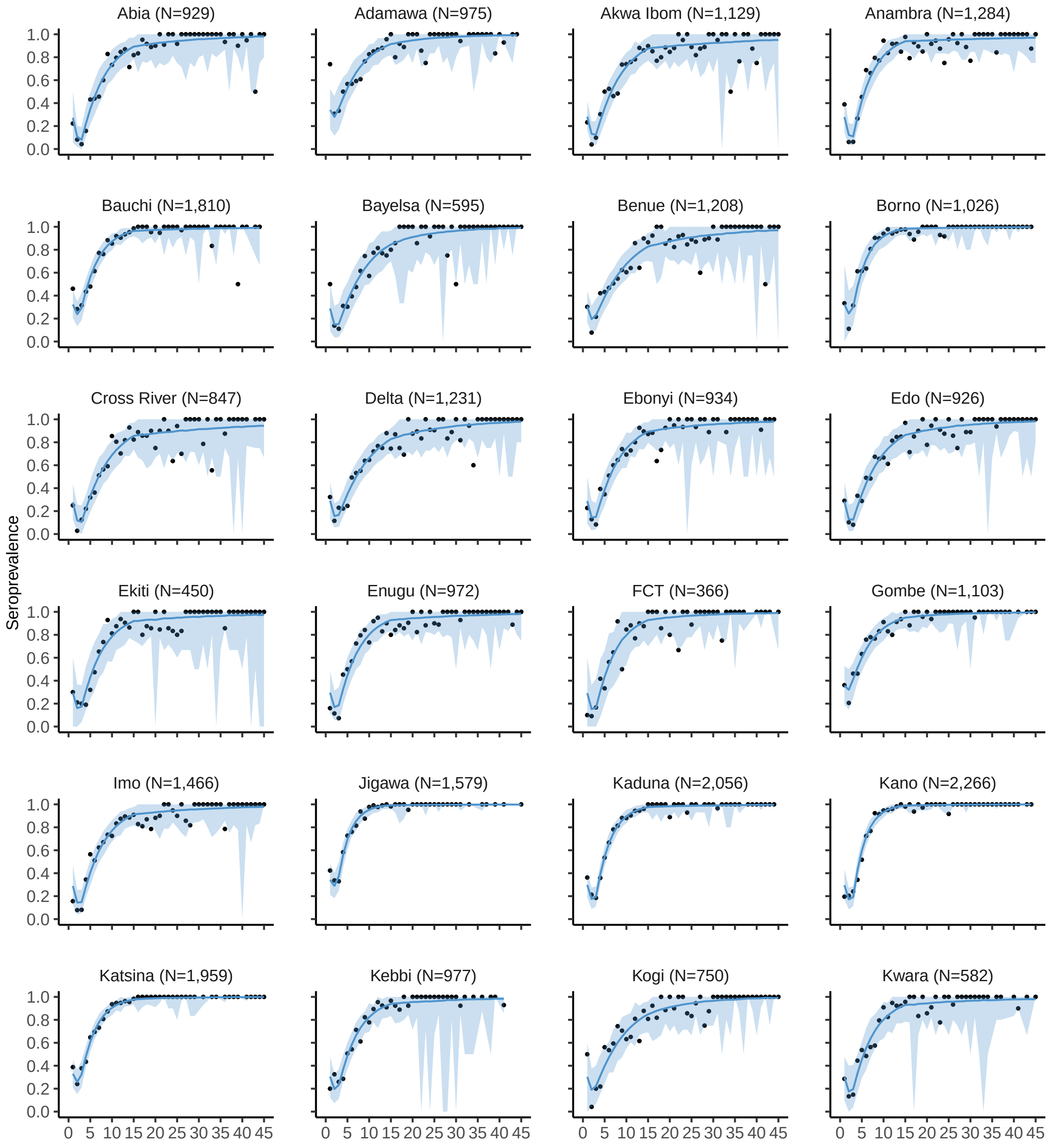
Abia (N=929)
Adamawa (N=975)
Akwa Ibom (N=1,129)
Anambra (N=1,284)
Bauchi (N=1,810)
Bayelsa (N=595)
Benue (N=1,208)
Borno (N=1,026)
Cross River (N=847)
Delta (N=1,231)
Ebonyi (N=934)
Edo (N=926)
Ekiti (N=450)
Enugu (N=972)
FCT (N=366)
Gombe (N=1,103)
Imo (N=1,466)
Jigawa (N=1,579)
Kaduna (N=2,056)
Kano (N=2,266)
Katsina (N=1,959)
Kebbi (N=977)
Kogi (N=750)
Kwara (N=582)
Seroprevalence
1.0
0.8
0.6
0.4
0.2
0.0
0 5 10 15 20 25 30 35 40 45

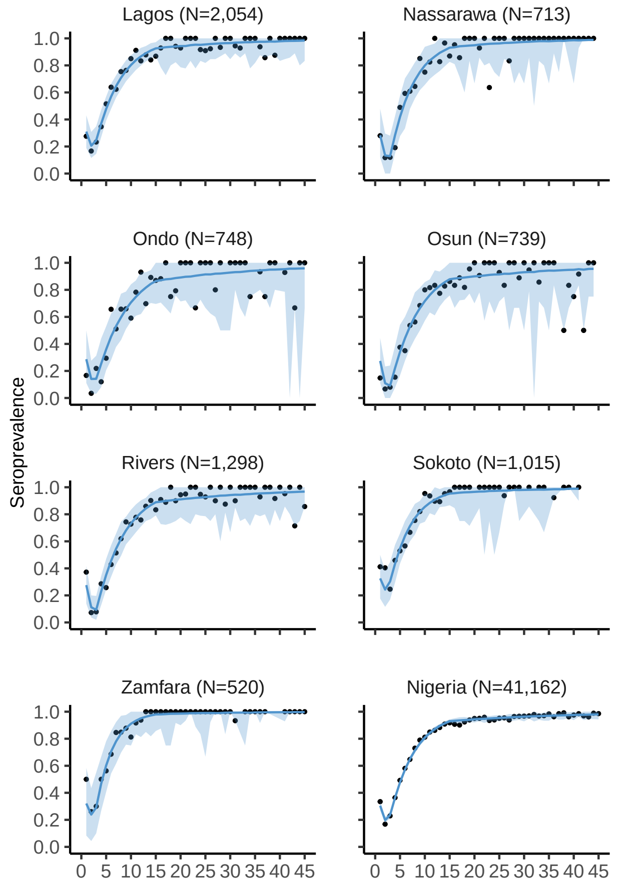

**Figure S1. Age-specific seroprevalence data for rubella with associated model fit.** The age-specific seroprevalence data at the state level ($N = 37$) and aggregated at the national level for sampled individuals aged 1 to 45 years are shown with the black circles. Posterior predictive checks for the model fit are shown with the blue line (median) and shaded area (90% credible interval).

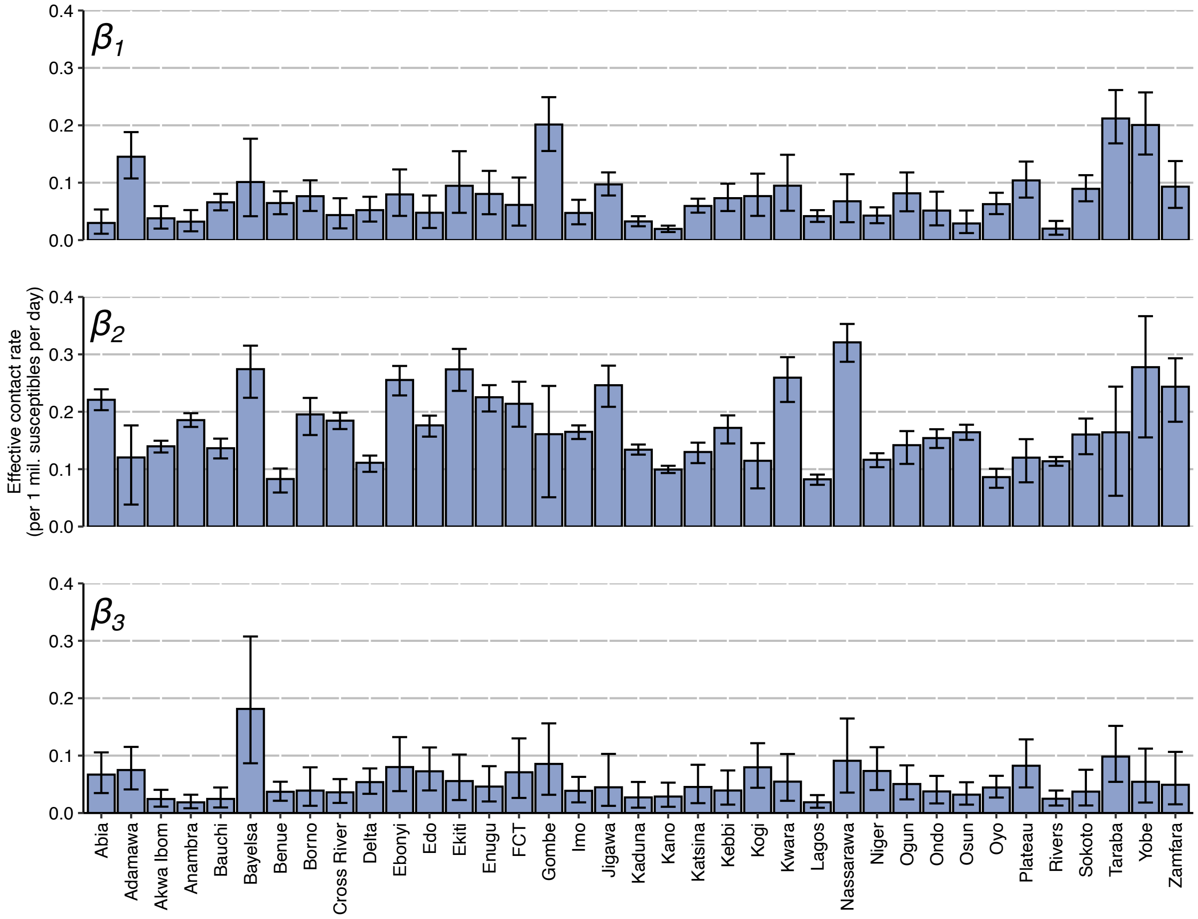

**Figure S2. Estimates of the effective contact rate among individuals in each state.** Mean and 90% credible interval of the effective contact rate (number of infectious contacts per 1 million susceptibles per day) for the three groups of individuals as defined in the WAIFW matrix $B$ (i.e. $\beta_1$, $\beta_2$ and $\beta_3$).

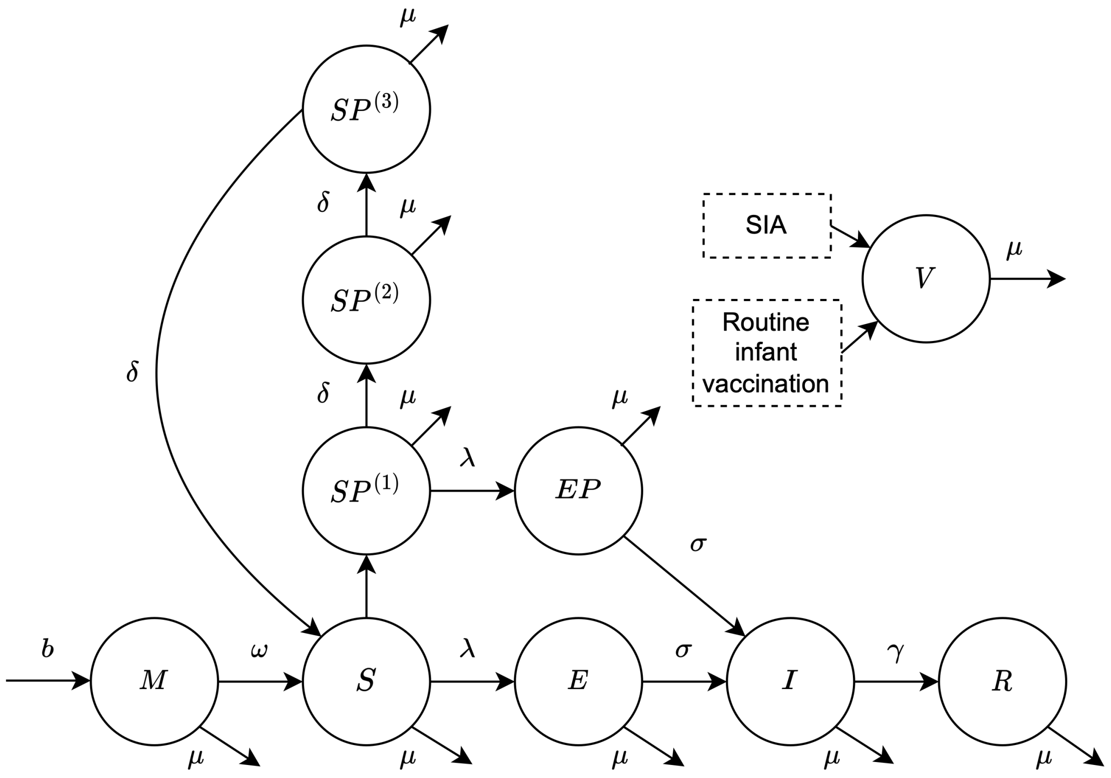

**Figure S3. Flow diagram of dynamic transmission model for a single age group.** Aging processes are not shown. The variables and parameters are described in Section 1.2 of the Supplementary Text. SIA=supplemental immunization activity.

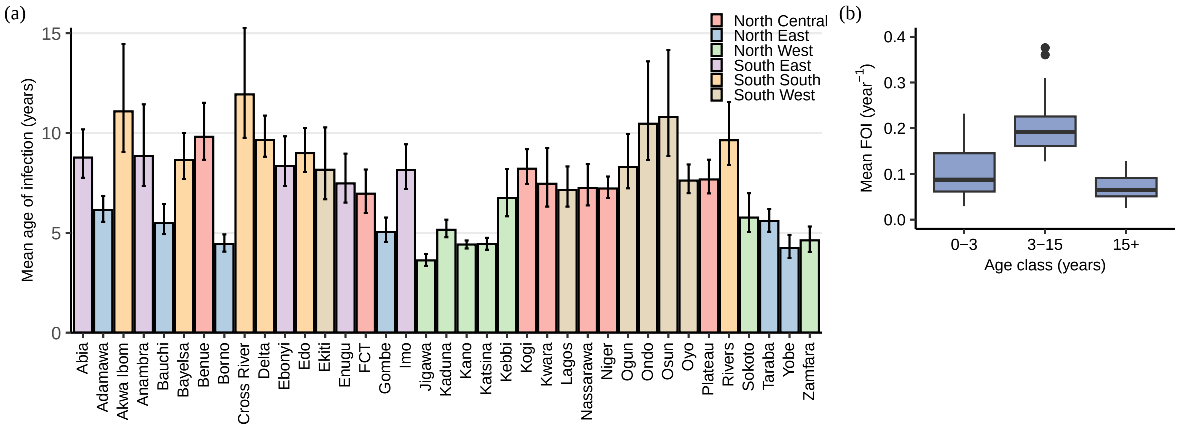

**Figure S4. Age-dependent rubella epidemiology across Nigeria. (a)** Estimated mean age of infection (years) in each state (mean and 90% credible interval). **(b)** The distribution of the mean estimates of the force of infection (FOI) for the three age groups (0-3 years, 3-15 years and 15+ years) across states.

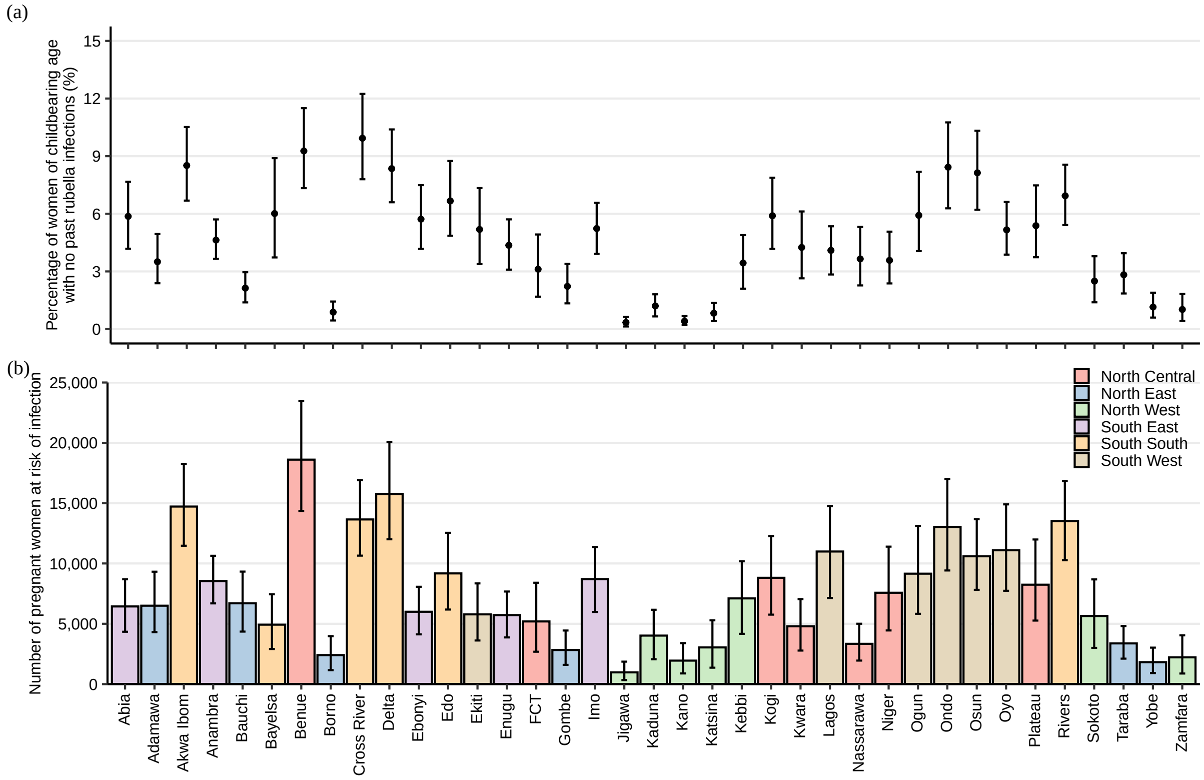

**Figure S5. Rubella epidemiology among women of reproductive age. (a)** State-level distributions of the predicted percentage of women of reproductive age (15-44 years) with no past rubella infections (mean and 90% credible interval). **(b)** State-level distributions of the estimated number of pregnant women at risk of rubella infection per year (mean and 90% credible interval).

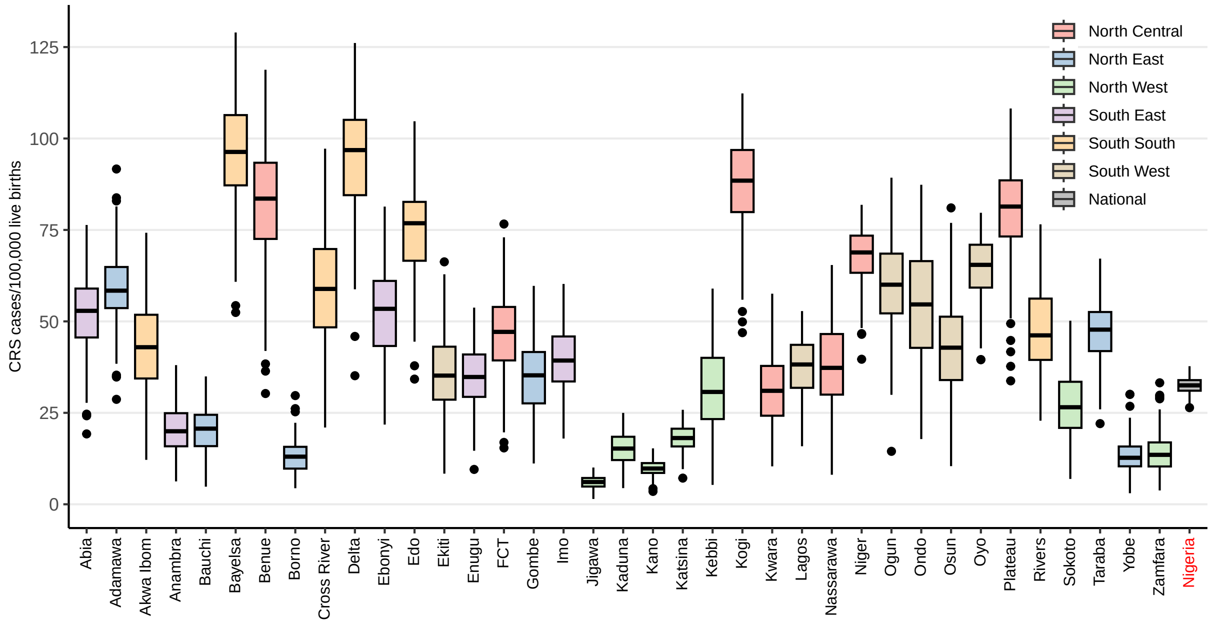

**Figure S6. Distribution of projected 30-year CRS burden in each state and at the national level in the absence of rubella vaccination.**

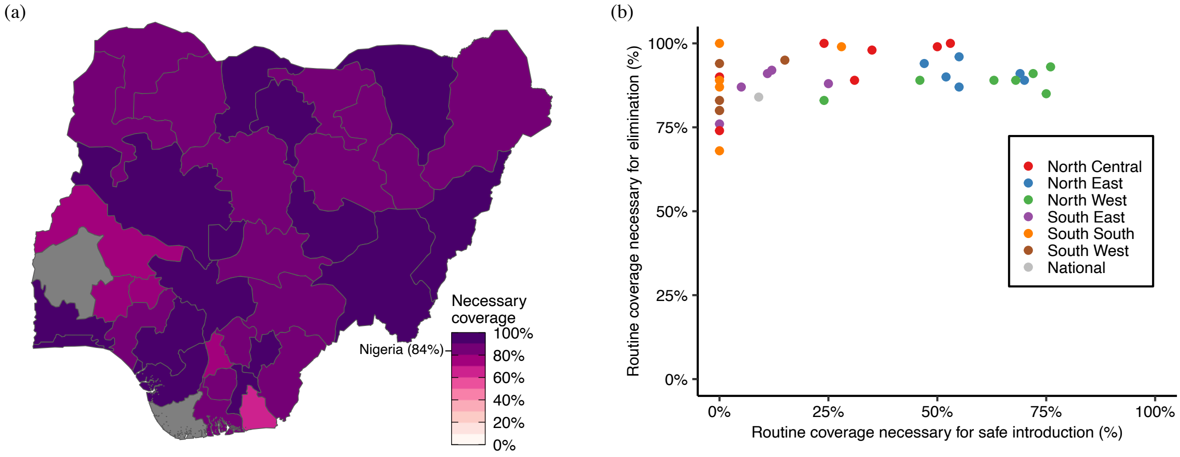

**Figure S7. Minimum necessary routine rubella vaccination coverage for CRS elimination. (a)** Map of minimum necessary routine rubella vaccine coverage for CRS elimination, defined as less than 1 CRS case per 100,000 live births during the 30th year. States that do not reach CRS elimination by the 30th year for any level of routine coverage (i.e. Oyo and Bayelsa) are colored grey. **(b)** Relationship between the minimum necessary routine coverage for safe rubella vaccine use (i.e. no relative increase in 30-year CRS burden) and the minimum necessary routine coverage for CRS elimination.

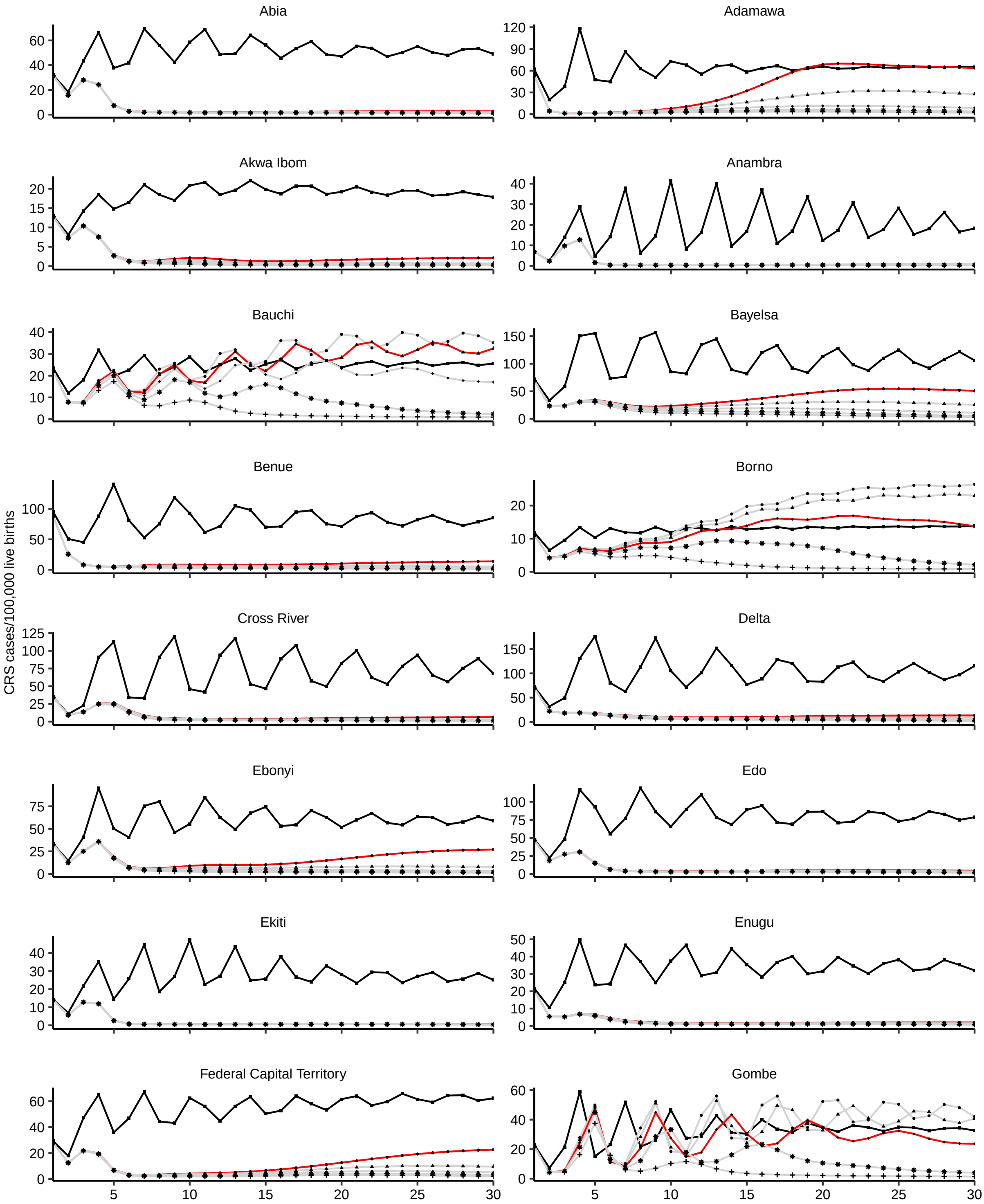

Abia
Adamawa
Akwa Ibom
Anambra
Bauchi
Bayelsa
Benue
Borno
Cross River
Delta
Ebonyi
Edo
Ekiti
Enugu
Federal Capital Territory
Gombe
CRS cases/100,000 live births

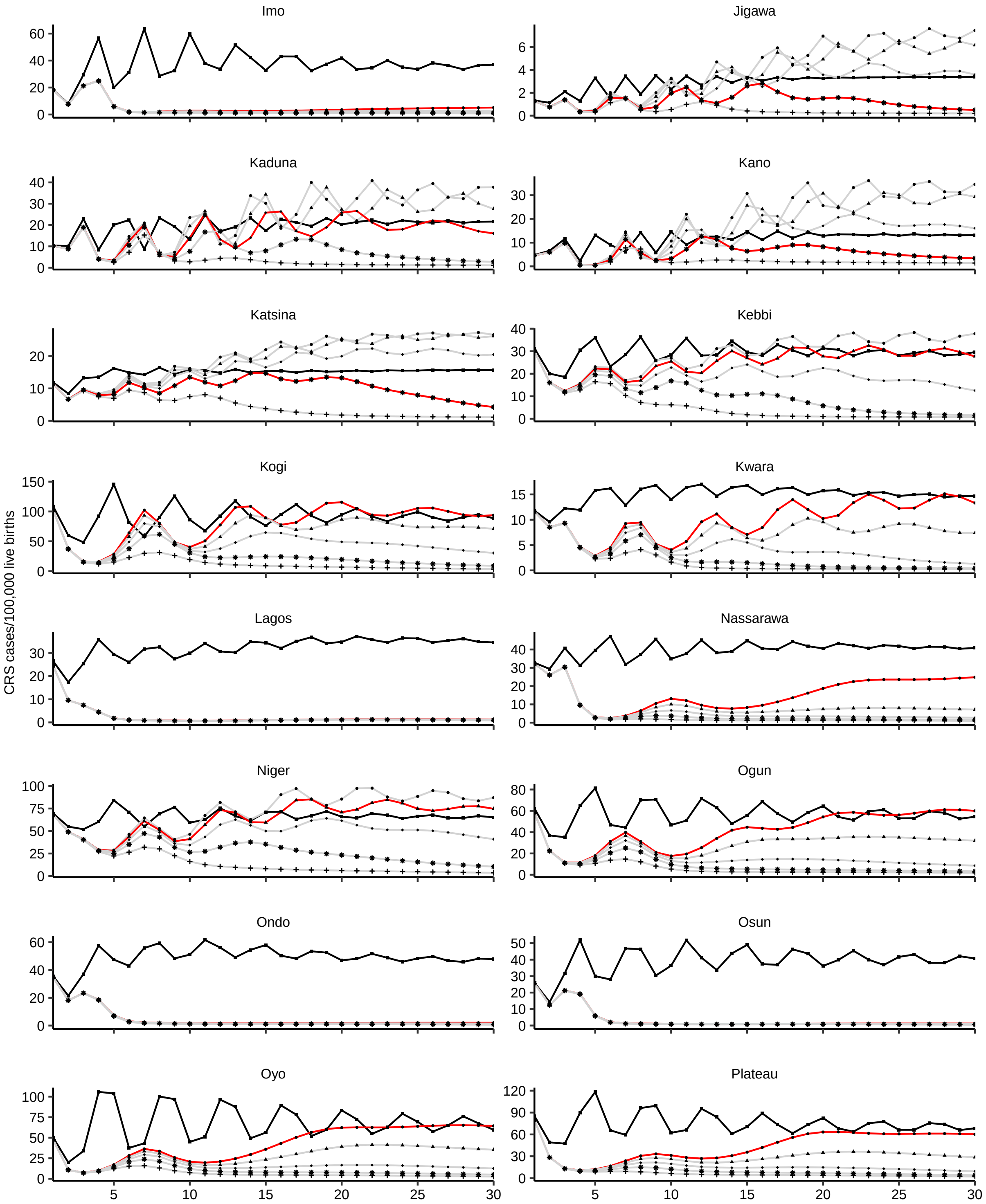
Imo
Jigawa
Kaduna
Kano
Katsina
Kebbi
Kogi
Kwara
Lagos
Nassarawa
Niger
Ogun
Ondo
Osun
Oyo
Plateau
CRS cases/100,000 live births
5
10
15
20
25
30

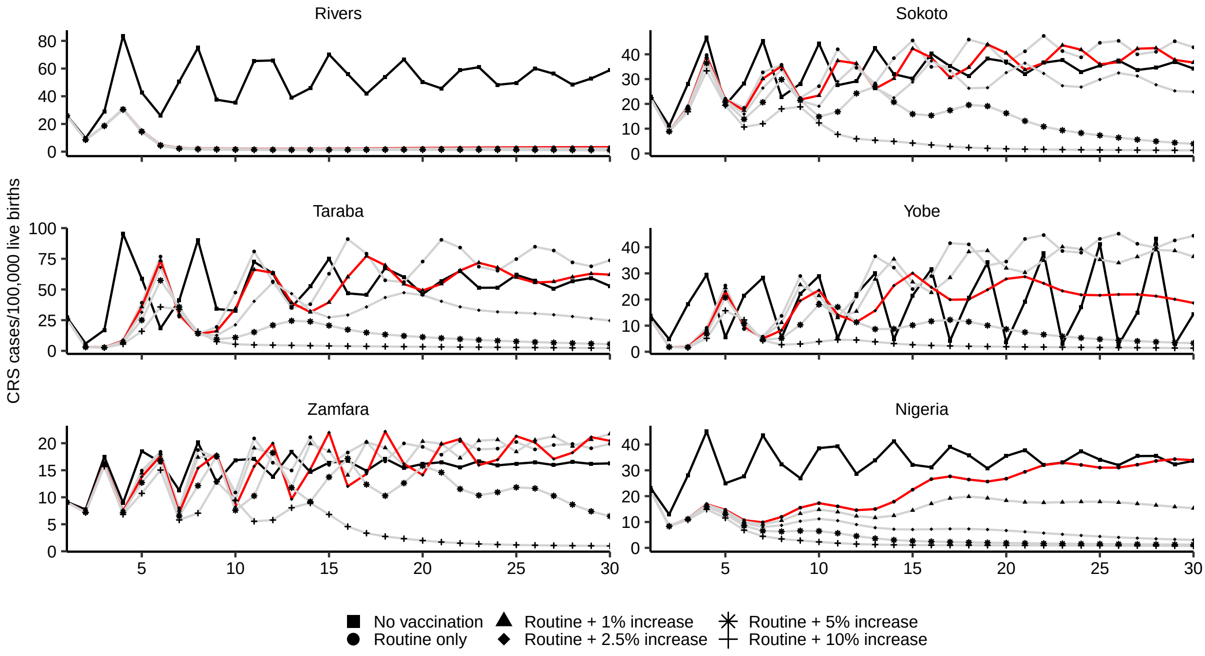

**Figure S8. Time series of yearly CRS incidence for different yearly rates of improvement in routine RCV coverage.** Estimated at the median $R_0$ in each state (black=no vaccination simulation; red=simulation for the minimum necessary rate of improvement in each state; grey=other simulations).

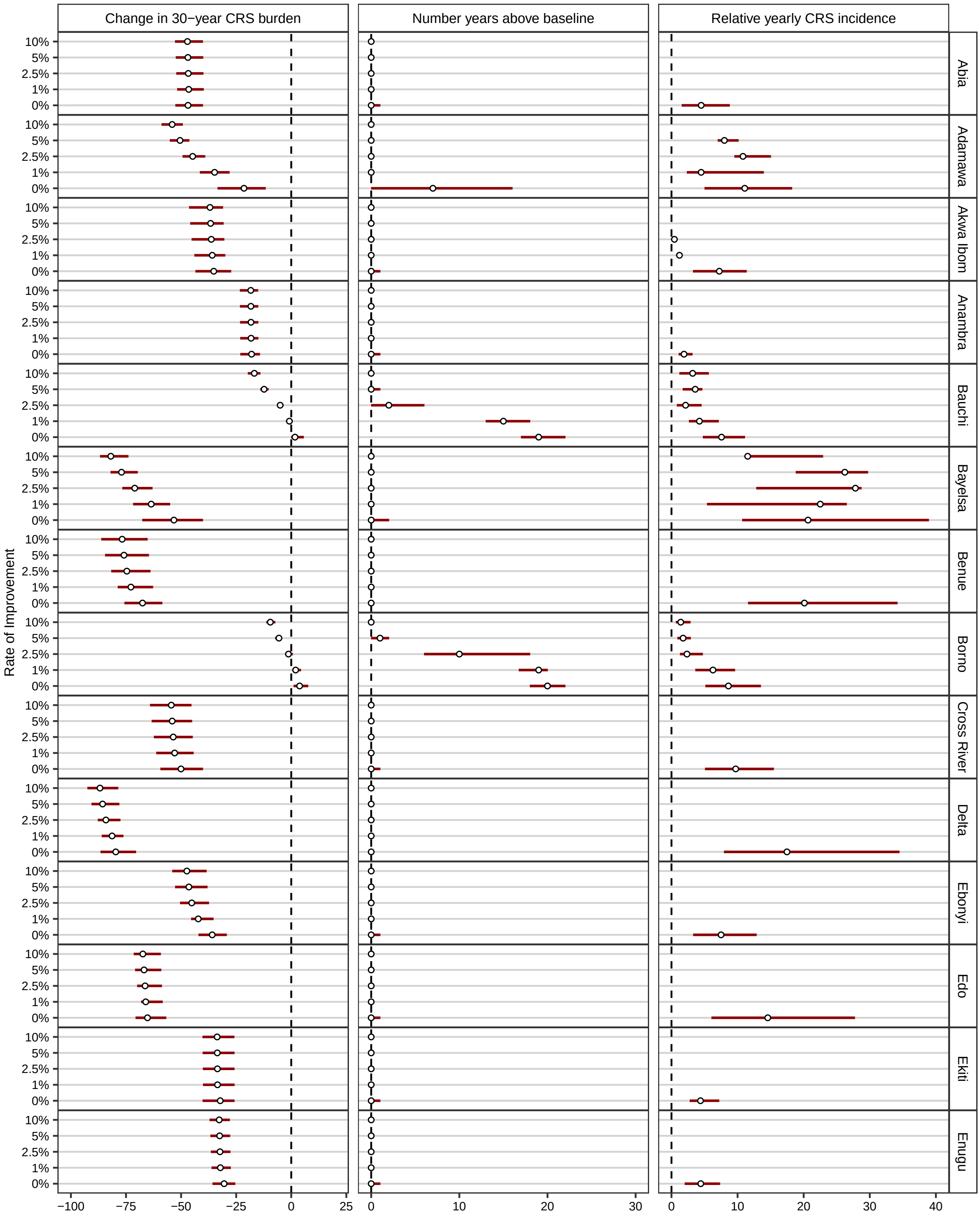
Change in 30−year CRS burden
Number years above baseline
Relative yearly CRS incidence
Rate of Improvement
10%
5%
2.5%
1%
0%
Abia
Adamawa
Akwa Ibom
Anambra
Bauchi
Bayelsa
Benue
Borno
Cross River
Delta
Ebonyi
Edo
Ekiti
Enugu
−100
−75
−50
−25
0
25
0
10
20
30
0
10
20
30
40

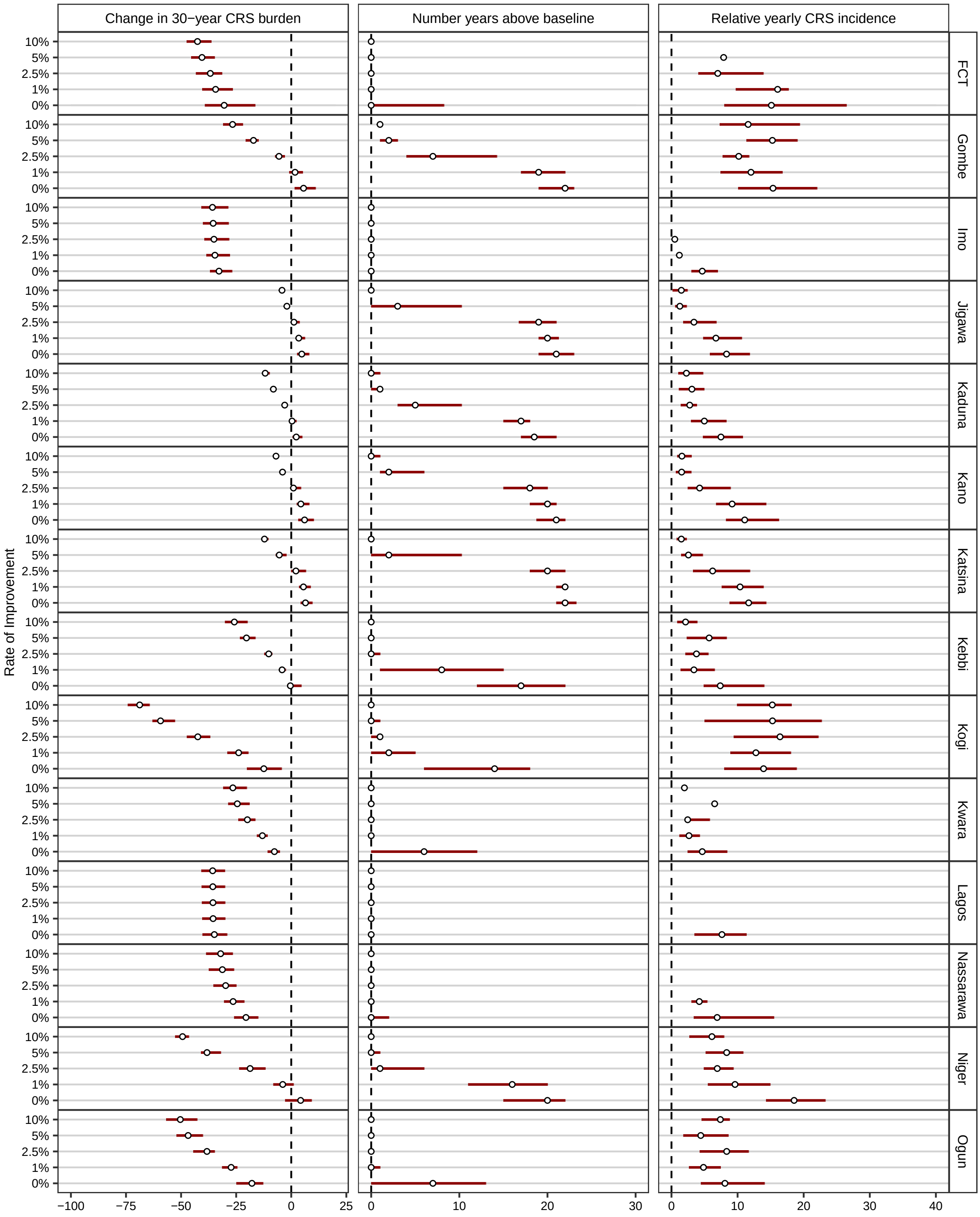
Change in 30−year CRS burden
Number years above baseline
Relative yearly CRS incidence
Rate of Improvement
10%
5%
2.5%
1%
0%
FCT
Gombe
Imo
Jigawa
Kaduna
Kano
Katsina
Kebbi
Kogi
Kwara
Lagos
Nassarawa
Niger
Ogun
−100
−75
−50
−25
0
25
0
10
20
30
0
10
20
30
40

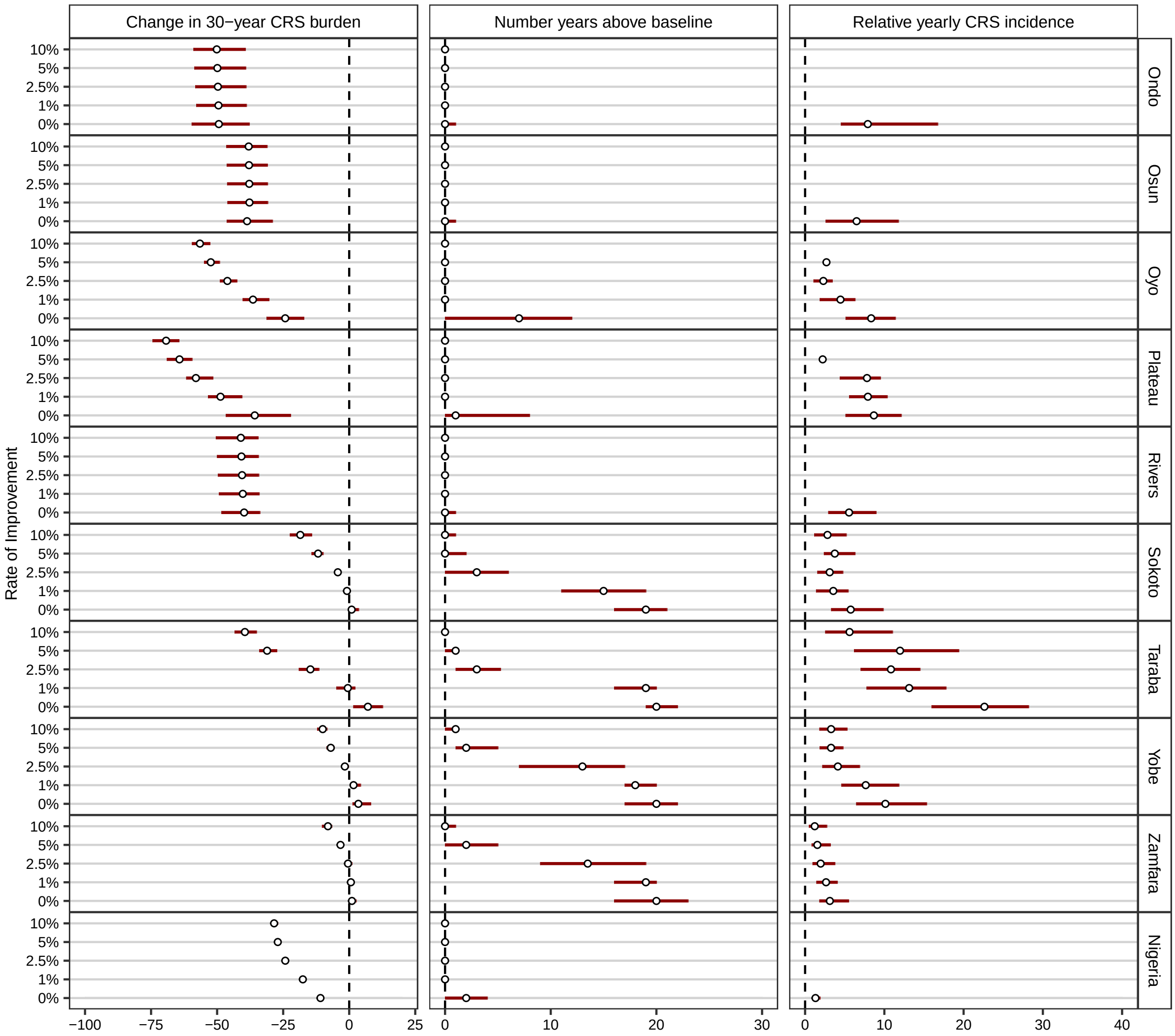

**Figure S9. Impact of routine RCV introduction on CRS burden for different yearly rates of improvement in coverage.** Change in 30-year CRS burden (CRS cases/100,000 live births) refers to the the change in 30-year CRS burden relative to pre-vaccination conditions after the introduction of routine vaccination for different rates of improvement in coverage. Number years above baseline and relative yearly CRS incidence correspond to the number of years CRS incidence was greater in the introduction scenarios than in the non-introduction scenario and the relative difference in the CRS incidence for those years, respectively. The median (open circle) and the interquartile range (red bar) are shown.

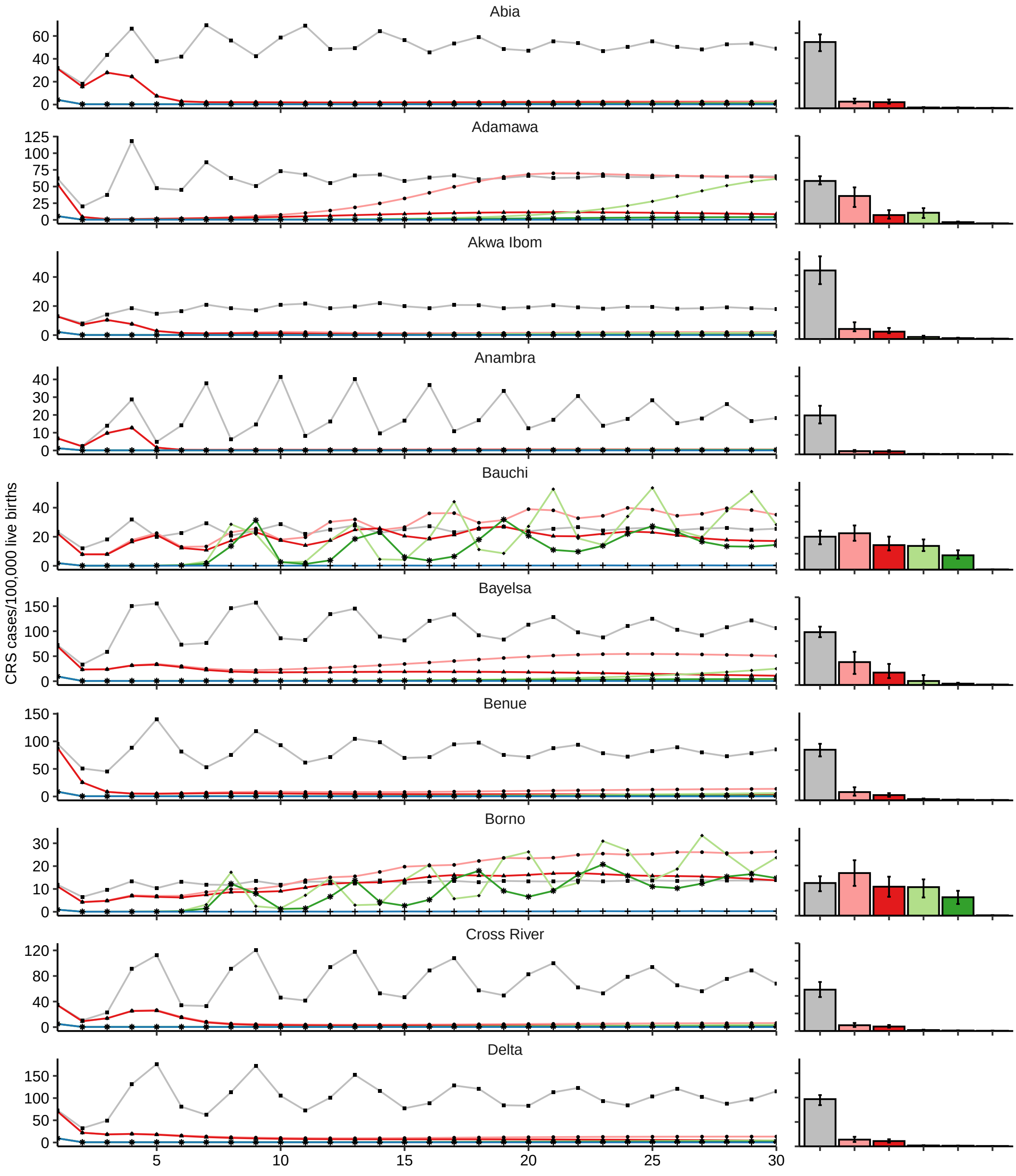
CRS cases/100,000 live births
Abia
Adamawa
Akwa Ibom
Anambra
Bauchi
Bayelsa
Benue
Borno
Cross River
Delta
5
10
15
20
25
30

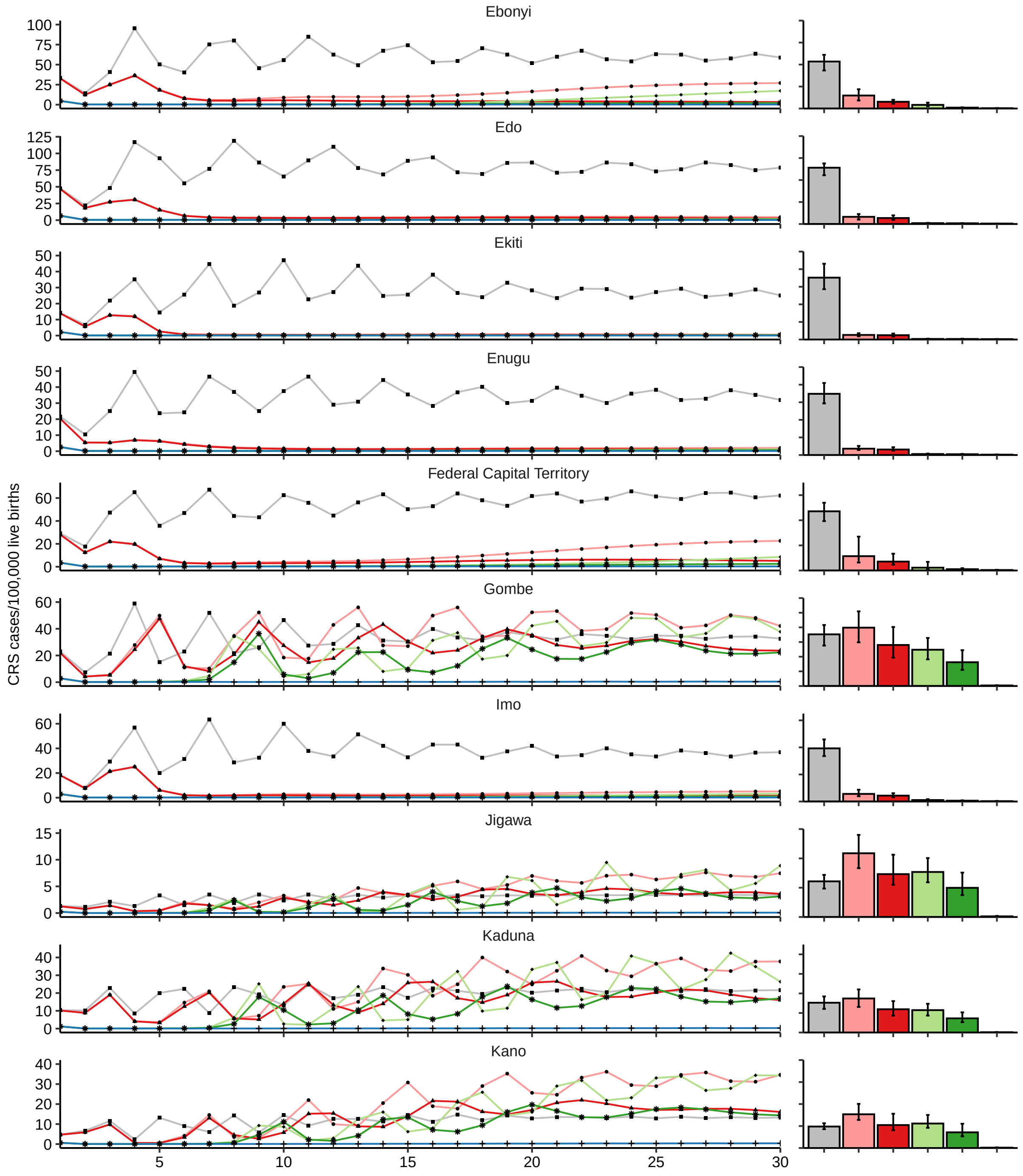
Ebonyi
Edo
Ekiti
Enugu
Federal Capital Territory
Gombe
Imo
Jigawa
Kaduna
Kano
CRS cases/100,000 live births

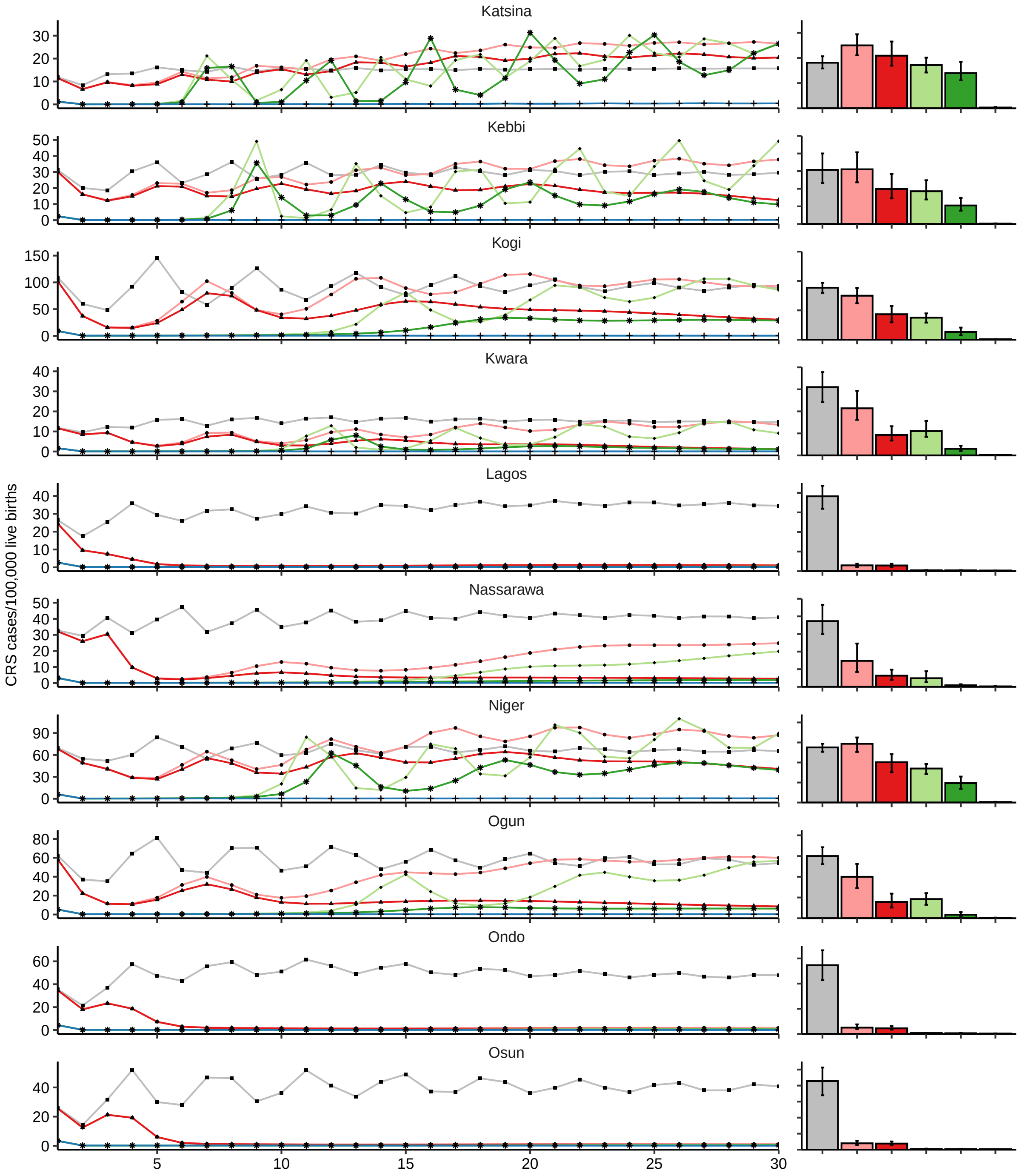
Katsina
Kebbi
Kogi
Kwara
Lagos
Nassarawa
Niger
Ogun
Ondo
Osun
CRS cases/100,000 live births
5
10
15
20
25
30

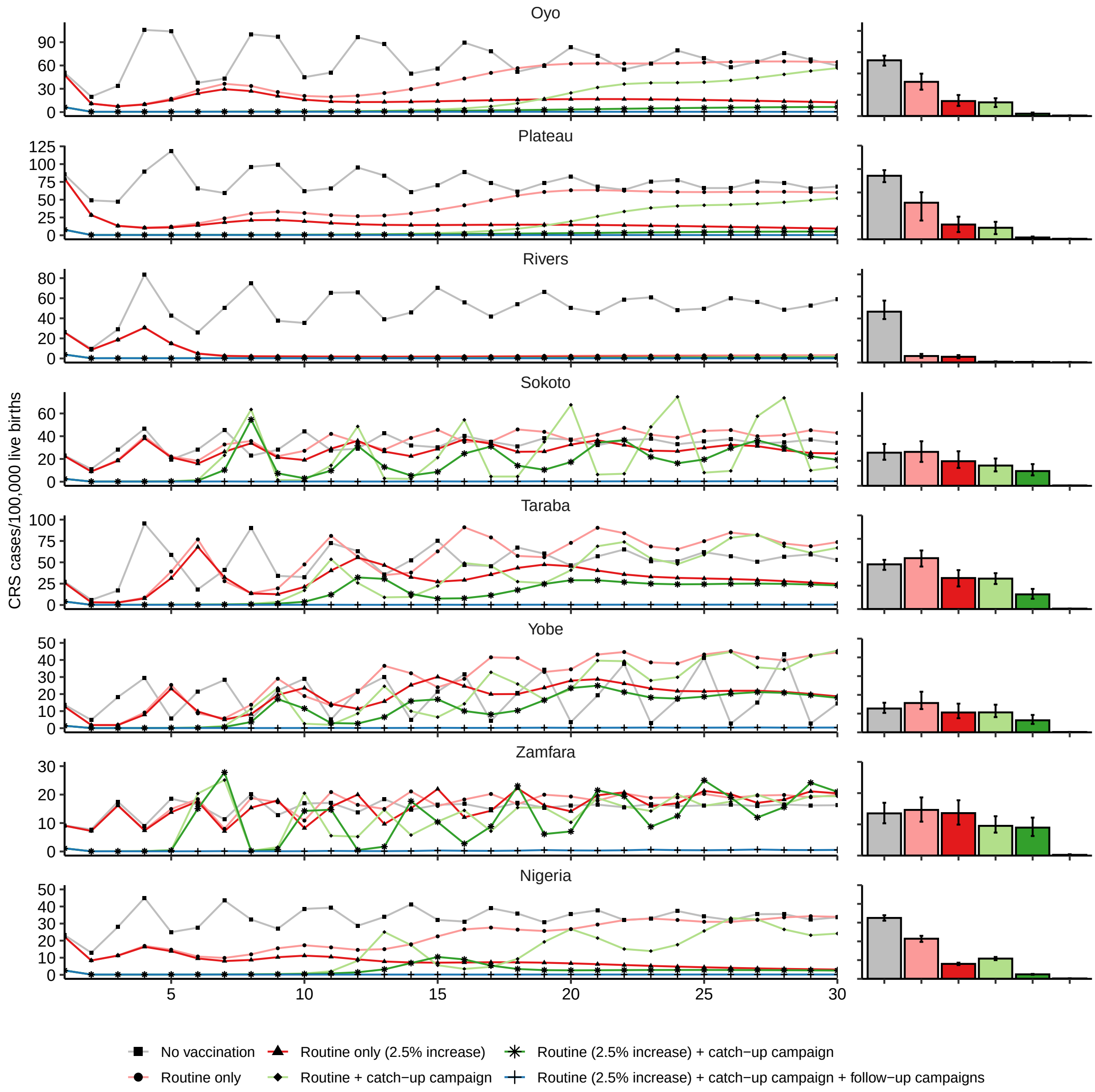

**Figure S10. Yearly CRS incidence time series and 30-year CRS burden under different vaccination scenarios.** The bar chart shows the distribution (median and interquartile range) of 30-year CRS burden (number of CRS cases/100,000 live births over 30 years) under each scenario.

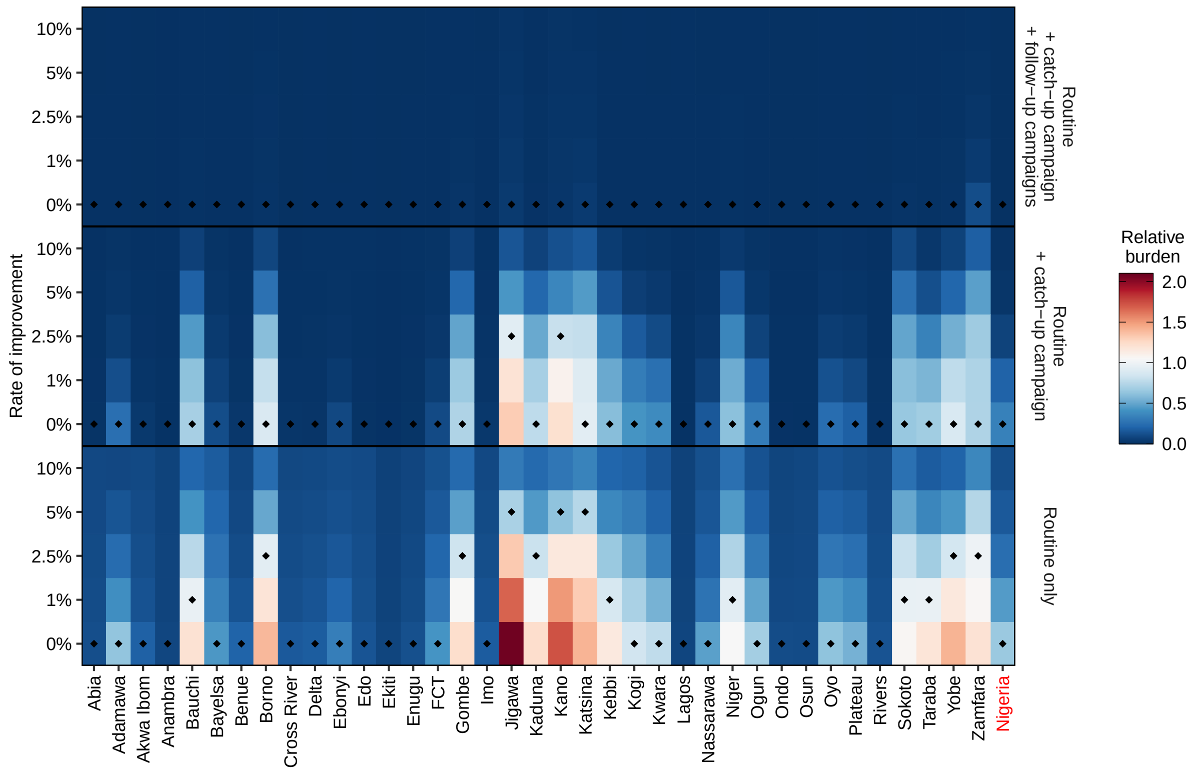

**Figure S11. Relative 30-year CRS burden to non-vaccination projections after introduction of RCV vaccination under all scenarios.** A relative burden of 1.5 indicates that the 30-year CRS burden is 50% greater in the introduction scenario compared to the non-vaccination projection. The minimum rate of improvement in routine RCV coverage necessary to have a reduction in 30-year CRS burden relative to pre-vaccination burden for each state under three different categories of scenarios (routine only, routine + catch-up campaign and routine + catch-up campaign + follow-up campaigns) are indicated by black triangles.

| Parameter | Distribution | Units | References |
|---|---|---|---|
| Force of infection ($\lambda_{i,j}$) | Lognormal($\mu = -2.6, \sigma = 0.7$) | 1/year | [12] |
| Average rate of loss of maternal passive immunity ($1/\omega$) | Lognormal($\mu = -1.04, \sigma = 0.74$) | year | [13] |

**Table S1. Weakly informative priors for the Bayesian MCMC estimation of the basic reproductive number in each state.** $\mu$ and $\sigma$ denotes the mean and standard deviation of the natural logarithm of each parameter. The same prior distribution was used for the force of infection in all three age groups and all states.

| State | MCV1 (%) | Minimum safe coverage (%) |
|---|---|---|
| Abia | 79 | 12 (0, 41) |
| Adamawa | 65 | 55 (33, 63) |
| Akwa Ibom | 64 | 0 (0, 0) |
| Anambra | 81 | 0 (0, 0) |
| Bauchi | 36 | 55 (31, 62) |
| Bayelsa | 71 | 27 (0, 47) |
| Benue | 64 | 0 (0, 0) |
| Borno | 46 | 70 (61, 78) |
| Cross River | 64 | 0 (0, 0) |
| Delta | 73 | 0 (0, 24) |
| Ebonyi | 64 | 11 (0, 40) |
| Edo | 81 | 28 (0, 46) |
| Ekiti | 86 | 0 (0, 49) |
| Enugu | 79 | 25 (0, 49) |
| FCT | 74 | 53 (33, 73) |
| Gombe | 29 | 52 (48, 70) |
| Imo | 71 | 5 (0, 45) |
| Jigawa | 56 | 75 (70, 85) |
| Kaduna | 43 | 63 (44, 73) |
| Kano | 56 | 76 (69, 84) |
| Katsina | 35 | 72 (61, 82) |
| Kebbi | 33 | 24 (0, 40) |
| Kogi | 45 | 24 (0, 40) |
| Kwara | 51 | 0 (0, 48) |
| Lagos | 90 | 0 (0, 51) |
| Nassarawa | 66 | 31 (0, 57) |
| Niger | 41 | 50 (23, 64) |
| Ogun | 52 | 15 (0, 40) |
| Ondo | 74 | 0 (0, 13) |
| Osun | 77 | 0 (0, 0) |
| Oyo | 62 | 32 (13, 46) |
| Plateau | 64 | 35 (0, 51) |
| Rivers | 73 | 0 (0, 9) |
| Sokoto | 19 | 46 (37, 63) |
| Taraba | 41 | 47 (39, 64) |
| Yobe | 46 | 69 (56, 75) |
| Zamfara | 12 | 68 (59, 78) |

**Table S2. Current MCV1 coverage and minimum safe levels of coverage for the introduction of routine rubella vaccination without campaigns.** MCV1 refers to the current measles-containing-vaccine first-dose coverage for each state. Median and 90% credible interval shown.

### References

1. Stan Development Team. RStan: the R interface to Stan. R package version 2.21.7. 2022. Available from: `https://mc-stan.org/`
2. Waaijenborg S, Hahné SJM, Mollema L, Smits GP, Berbers GAM, Klis FRM van der, et al. Waning of Maternal Antibodies Against Measles, Mumps, Rubella, and Varicella in Communities With Contrasting Vaccination Coverage. The Journal of Infectious Diseases 2013 May; 208:10–16
3. Stan Development Team. Stan Modeling Language Users Guide and Reference Manual. Version 2.31. 2022. Available from: `https://mc-stan.org/`
4. Anderson RM and May RM. Age-related changes in the rate of disease transmission: implications for the design of vaccination programmes. eng. The Journal of hygiene 1985; 94:365–436
5. Patel MK, Antoni S, Danovaro-Holliday MC, Desai S, Gacic-Dobo M, Nedelec Y, et al. The epidemiology of rubella, 2007–18: an ecological analysis of surveillance data. eng. The Lancet global health 2020; 8:e1399–e1407
6. United Nations, Department of Economic and Social Affairs, Population Division. Abridged Life Table - Both Sexes. 2019. Available from: `https://population.un.org/wpp/Download/Files/5_Archive/WPP2019-Excel-files.zip`
7. Farrington CP, Kanaan MN, and Gay NJ. Estimation of the basic reproduction number for infectious diseases from age-stratified serological survey data. eng. Journal of the Royal Statistical Society. Journal of the Royal Statistical Society Series C 2001; 50:251–92
8. Diekmann O, Heesterbeek J, and Metz J. On the definition and the computation of the basic reproduction ratio R 0 in models for infectious diseases in heterogeneous populations. eng. Journal of mathematical biology 1990; 28
9. Metcalf CJE, Lessler J, Klepac P, Cuts F, and Grenfell BT. Impact of birth rate, seasonality and transmission rate on minimum levels of coverage needed for rubella vaccination. Epidemiology and Infection 2012; 140:2290–301
10. Powell MJD. A Direct Search Optimization Method That Models the Objective and Constraint Functions by Linear Interpolation. *Advances in Optimization and Numerical Analysis*. Ed. by Gomez S and Hennart JP. Dordrecht: Springer Netherlands, 1994 :51–67
11. Johnson SG. The NLopt nonlinear-optimization package. 2023. Available from: `https://nlopt.readthedocs.io/en/latest/`
12. Farrington CP. Modelling forces of infection for measles, mumps and rubella. eng. Statistics in medicine 1990; 9:953–67

13. Nicoara C, Zäch K, Trachsel D, Germann D, and Matter L. Decay of Passively Acquired Maternal Antibodies against Measles, Mumps, and Rubella Viruses. eng. Clinical and Diagnostic Laboratory Immunology 1999; 6:868–71
